# Impact of Digital Contact Tracing on Pandemic Control Analysed with Behaviour-driven Agent-based Modelling

**DOI:** 10.1101/2025.01.20.25320711

**Authors:** Luis Ignacio Lopera González, Göran Köber, Göran Kirchner, Justus Benzler, Oliver Amft

## Abstract

We disentangle the efficacy of individual non-pharmaceutical interventions (NPIs), including digital contact tracing (DCT), with a novel behaviour-driven agent-based model (ABM) to inform ongoing pandemic preparedness efforts. Our model’s Zeitgeber architecture delineates contextual characteristics, including daytime, daily routines, locations, and activities. Our method determines each agent’s current location and behaviour in a realistic environment under the restrictions of NPIs. We model viral load transfer between agents from contact duration, distance, and the infected agent’s infectiousness level. We examine the effects of DCT adoption, adherence, and compliance, both individually and combined with other NPIs, on key pandemic indicators, thus providing novel insight into infection dynamics. DCT combined with other NPIs reduces the total infections up to 52% for realistic behaviour, whereas DCT alone yielded a 43% reduction. Surprisingly however, some NPI combinations do not improve pandemic parameters. Our approach offers fine-grained analysis capabilities on the effectiveness of NPI combinations that cannot be obtained in human studies due to confounding effects. Thus our approach can inform future pandemic control efforts and prioritisation in pandemic preparedness.

## Introduction

Upon the global outbreak of coronavirus SARS-CoV-2, digital contact tracing (DCT) applications (“apps”) were introduced by health authorities worldwide for the first time to help end infection chains. For example, in 2021, TraceTogether (a DCT app administered in an opt-out scheme) was used by more than 90% of Singaporeans^1^. DCT is intended to avoid other, more disruptive non-pharmaceutical interventions (NPIs), e.g., lockdowns and generalised strict social distancing^2^.

With DCT effectiveness, we refer to reducing the number of infected individuals and flattening the curve of infections^3^. By design, DCT could complement the comparably slow, laborious, and error-prone manual contact person tracing (MCT) of an infected individual’s social contact persons. MCT is usually done by health authorities. Instead, smartphone DCT apps leverage the device’s autonomous monitoring and analysis of Bluetooth signals received from other nearby smartphones and their attenuation. Thus, DCT apps can estimate contact duration and distance to other smartphones. While DCT is a sustainable NPI that can reduce infections during a pandemic, a rigorous evaluation is needed to assess its public-health benefits, weigh benefits against unwanted effects, including costs, inform the public, and guide DCT app development into an ethical and accepted tool for future outbreaks of infectious diseases^4^.

Several studies and reviews have explored the effectiveness of DCT^2–9^, with experimental results showing mixed outcomes, ranging from effective^10–12^ to ineffective^5,13^. However, evaluating the impact of DCT empirically with observational data is infeasible due to overlapping NPI effects during the COVID-19 pandemic. Thus, comprehensive individual-level simulations are warranted and may help to identify optimal combinations of DCT and other NPIs for pandemic preparedness.

Various approaches have been proposed to model and simulate contact tracing mechanisms in a pandemic. Compartmental models are used to model the pandemic continuum based on estimated epidemiological parameters^14,15^. Usually, compartmental models presuppose homogeneity^16^, i.e., people are interchangeable. However, from an epidemiological perspective, the unique social behaviour of individuals significantly influences virus transmission. In this context, the compliance of highly interconnected individuals (i.e., potential superspreaders) with NPIs is more critical than that of individuals with fewer social connections^17^. Agent-based models (ABMs) can naturally encode behaviour complexity and thus deal with heterogeneous social structures^18^. Additionally, ABMs allow modellers to specify contact networks, determining agent contacts, and the probability of virus propagation^9^. Nodes in a contact network represent individuals and the network edges their interaction. Existing ABMs deploy predefined contact networks for access-restricted spaces, e.g., home or office, or use randomly created networks for public spaces^19^. Thus, current ABMs summarise interactions between individuals while abstracting the actual diversity of individual interactions (e.g., contact duration) and their dynamic character (e.g., contacts due to individual behaviour patterns). Current ABMs typically encode NPIs as changes in the network topology, either by removing edges or by changing the edges’ transmission probability prior to the simulation^8^. The effect of an NPI needs to be known or hypothesised to modify an ABM’s internal networks^20^.

In this paper, we utilise ABMs to model the nuances of individual behaviour, allowing meso-level^21^ entities (e.g., contact networks) and quantities (e.g., location of infections) to emerge, in addition to macro-level quantities (e.g., wave durations). Our approach contrasts with traditional ABM methods that predefine meso-level quantities and investigate emerging macro-level quantities. In what we refer to as behaviour-driven ABMs, behaviour and social interactions are no longer assumed to be homogeneous across a population but depend on each individual’s actions and opportunities provided by their facet of a virtual world. In our simulation, activities are facilitated by individual needs, locations, health status, other individuals, and, crucially, NPIs. As an individual’s behaviour determines their social interactions, correspondingly, behaviour is affected by preceding social interactions (e.g., to quarantine). Consequently, in a behaviour-driven ABM, the behaviour patterns of individuals cannot be precomputed.

In our approach, behaviour is represented by parameterisable statistical models of contextual characteristics: daytime, location, and activities^22^ that are configured based on the agent’s specific profile, daytime, and location. The simulation is controlled by a Zeitgeber that determines how agents react to individual necessities and their environment, e.g., sleep schedule and bed availability to sleep^23^. Activities are generated as instances with start and end on the timeline. In our behaviour model, an NPI is equivalent to an extrinsic input that modifies an individual’s behaviour and thus the activities. For example, upon receiving a contact notification, an agent may decide to stay at home. Similarly, closing bars and restaurants inhibits eating-out activities and, thus, contact opportunities.

Our pathogen model is finetuned on SARS-CoV-2 and comprises a time-dependent infectiousness level for every agent, a distance-dependent viral load transfer function, and a wash-out, i.e., viral load removal function. Upon a contact between an infectious agent and a susceptible agent, a viral load is transferred that depends on agent distance, contact duration, and the infectious agent’s infectiousness level. The transferred viral load is added to the susceptible agent’s viral load level. If the viral load level is not “washed-out” by the removal function before further viral load is accumulated, the agent’s viral load level may reach a critical threshold, upon which the agent will transition from exposed to infected state (i.e., a self-sustained viral state). Fig. 1 provides an overview on the Zeitgeber, pathogen model, and examples of the virtual world infrastructure.

**Fig. 1:**
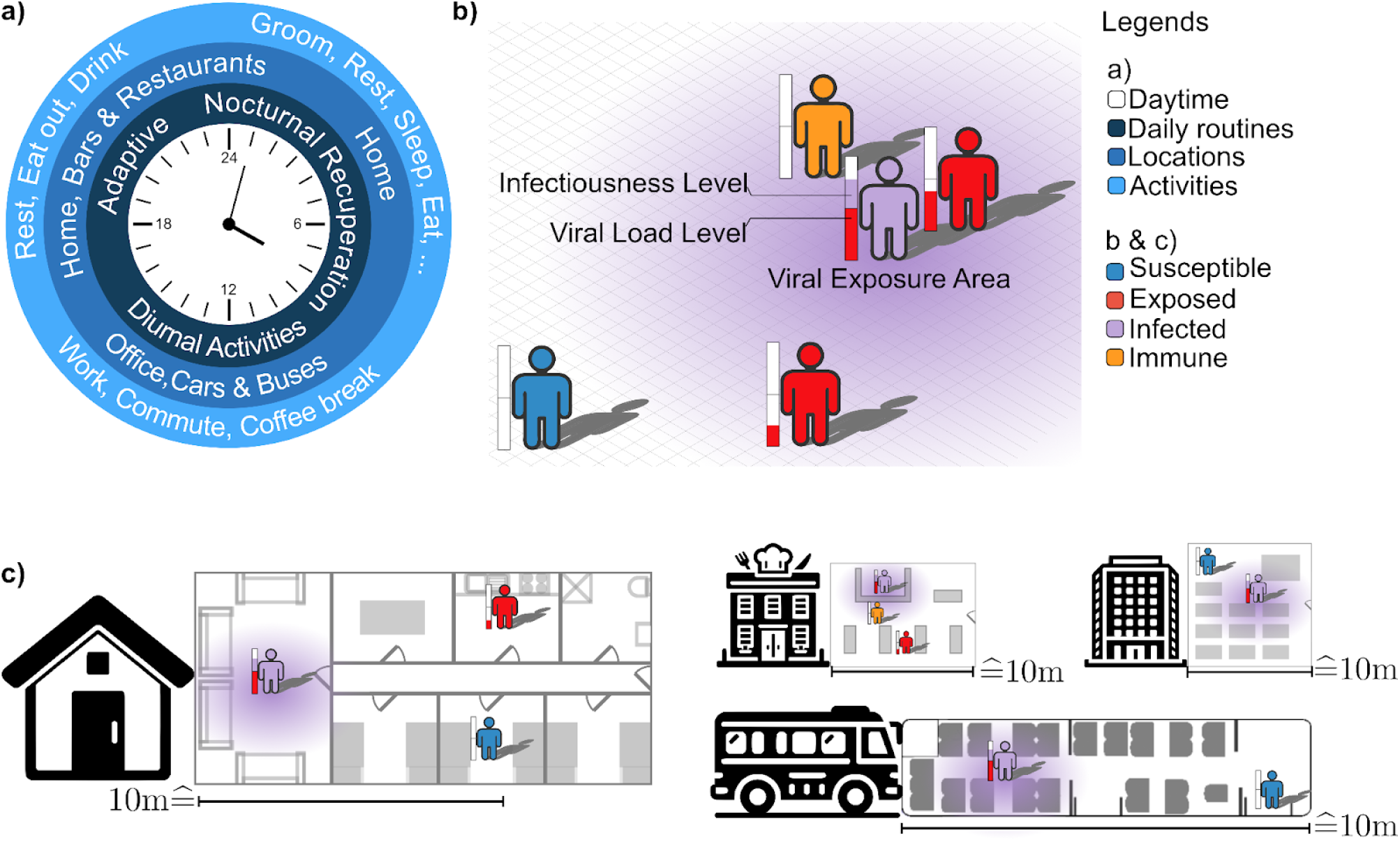
Overview of the behaviour-driven ABM approach. (a) Zeitgeber architecture delineates contextual characteristics: daytime, daily routines, locations, and activities. Simulation timekeeping controls the agents’ locations and location-based activities that result in contacts (see Fig. 5 for details). (b) The pathogen model to describe an agent-specific dynamically accumulated viral load level due to contact duration and distance between susceptible and infectious agents and a time-dependent wash-out. The transition from exposed to infected (i.e., self-sustaining viral state) is denoted by a critical threshold on the viral load level. An agent’s time-dependent infectiousness level influences disease progression (see Tab. 1 for disease states and characteristics). (c) Snapshots of virtual world infrastructure and agent instances, illustrating the spatial and dynamic character of viral load level (susceptible agents) and viral exposure area (infected agents). Details of all submodels can be found in the Methods section. An example simulation video can be found in Supplementary material.

**Table 1:**
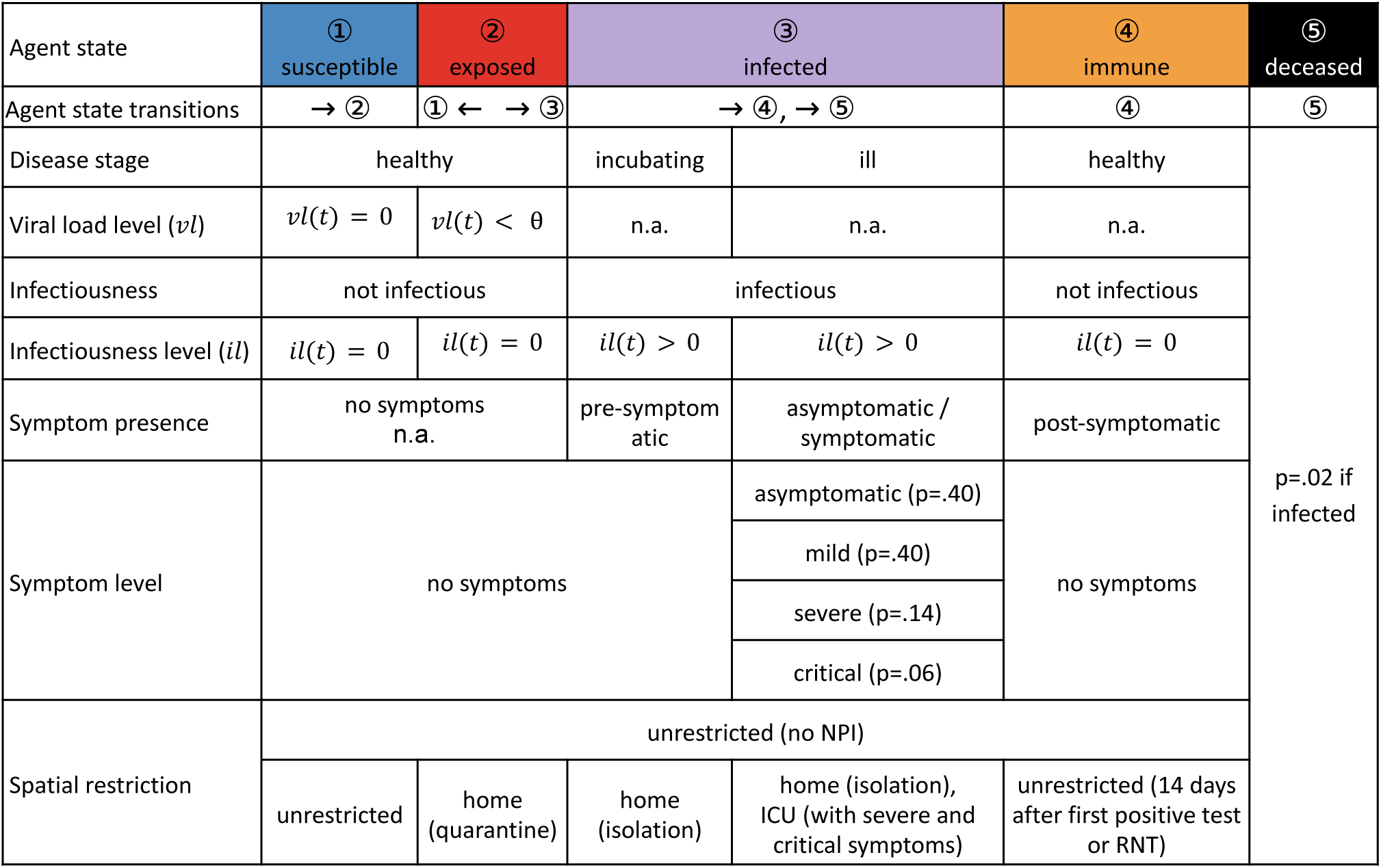
Terminology overview and key pathogen model concepts.

DCT seeks to inhibit virus transmission by detecting an infected individual’s risk contacts and changing the contact persons’ behaviour and their further contact opportunities via quarantine. For a DCT app to be effective, (1) it needs to be downloaded and activated, e.g., a user registers and activates Bluetooth (termed DCT adoption^24^); (2) the app user shall notify others by sharing a positive testing result (termed DCT adherence^25^); and (3) the app user shall follow recommendations upon receiving an app notification (termed DCT compliance^26^). Both, simulation and experimental analyses have shown that DCT adoption, even at or below 50%, may help in a containment strategy^27,28^. However, investigations suggest that adherence and compliance affect DCT effectiveness too^29^. Box 1 lists all considered NPIs.

#### Non-pharmaceutical interventions (NPIs) covered in this work Contact tracing

##### Informal contact person tracing (ICT)

Agents who test positive inform other (known) agents that were exposed to them. The decision to inform a contact person is based on the type of relationship with that person.

##### Manual contact person tracing (MCT)

Contact tracers interview agents who tested positive about other (known) agents that were earlier exposed to them. Contact persons are then informed based on the type of inter-agent relationships.

##### Digital contact tracing (DCT)

A proportion of the population is equipped with a DCT app that records duration and distance to other individuals equipped with the DCT app. The DCT app warns its users if there was a possible exposure to an individual who tested positive. Literature sources use “digital proximity tracing” synonymously with DCT.

##### DCT-related terminology

“DCT adoption” – proportion of people, who downloaded and activated, e.g., registered in the app and activated Bluetooth; “DCT adherence” – proportion of app users, who notify others by sharing a positive test result; “DCT compliance” – proportion of app users, who follow recommendations upon receiving an app notification.

###### Default intervention

As a simulation default, infected agents isolate after a positive test and may return as recovered agents after 14 days. In addition, we apply ICT and MCT (described above) in all simulations.

###### Contact restrictions and confinement relaxations

Contact restrictions may confine (i.e. quarantine or isolate) agents to their homes.

##### Quarantine contact person household (QCH)

With QCH, all household members of an agent, who is identified as contact person of an infectious agent, are quarantined for 14 days. RNT can shorten the quarantine period, as described above. Furthermore, QCH quarantines all household members of an infected agent.

##### Closure of bars and restaurants (CBR)

We consider bars and restaurants as public places that are inaccessible under CBR.

##### Require negative test to exit (RNT)

RNT reduces the default 14-day quarantine and isolation period by releasing agents based on a negative test. We assume that if an agent is isolated or in quarantine, the agent will get tested. For infected agents, the test happens once they recover, and for susceptible agents, the test happens five days after the start of the quarantine. It takes one additional day for the test to be processed and its result to be received by the agent. Upon a negative test result, agents are permitted to resume their normal activities. Thus, RNT is always used in combination with another NPI.

#### Definition of pandemic characteristics

Pandemic characteristics are emergent quantities measured on the virtual population. The metrics are used to validate and measure the effect of the simulated NPIs.

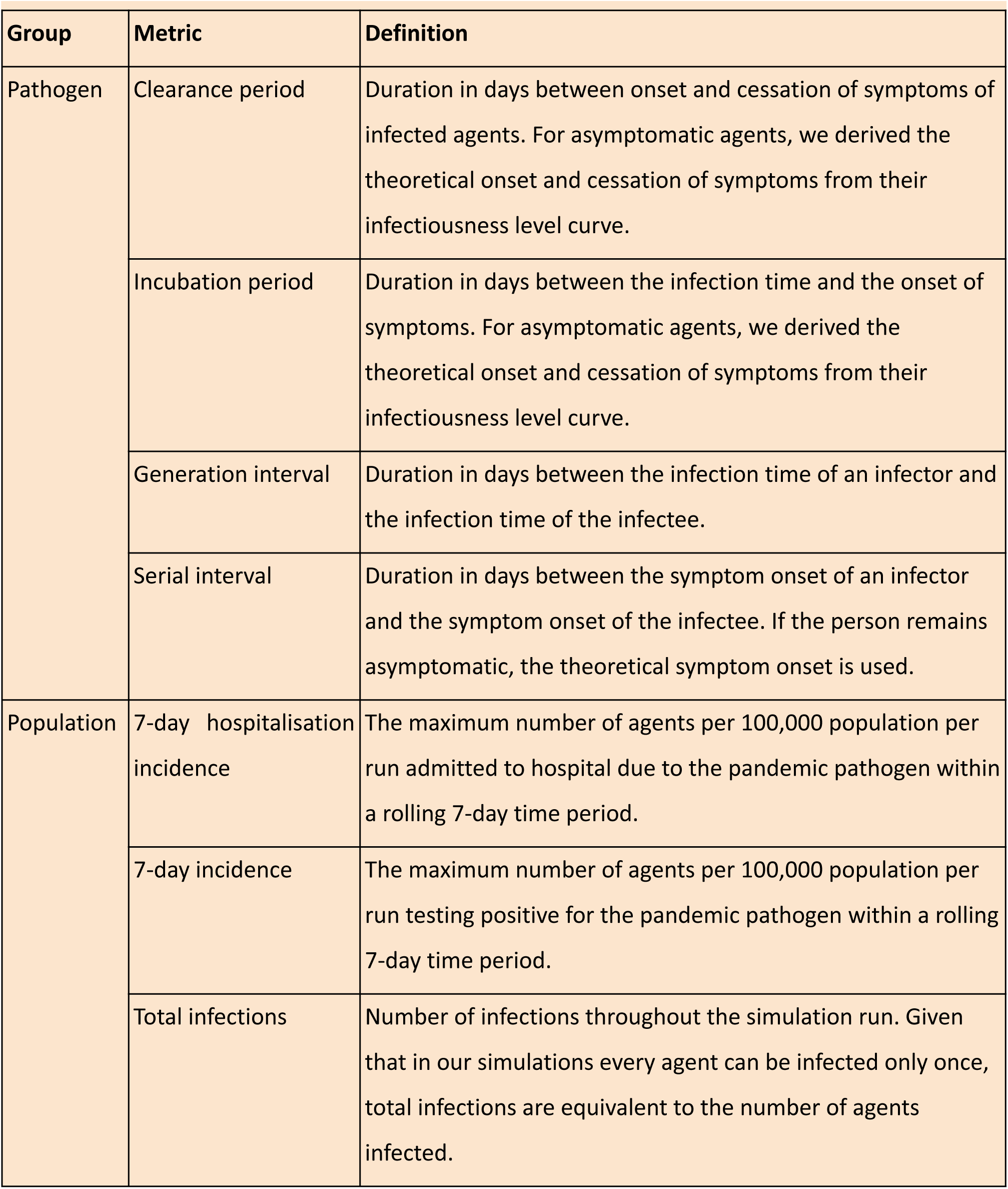
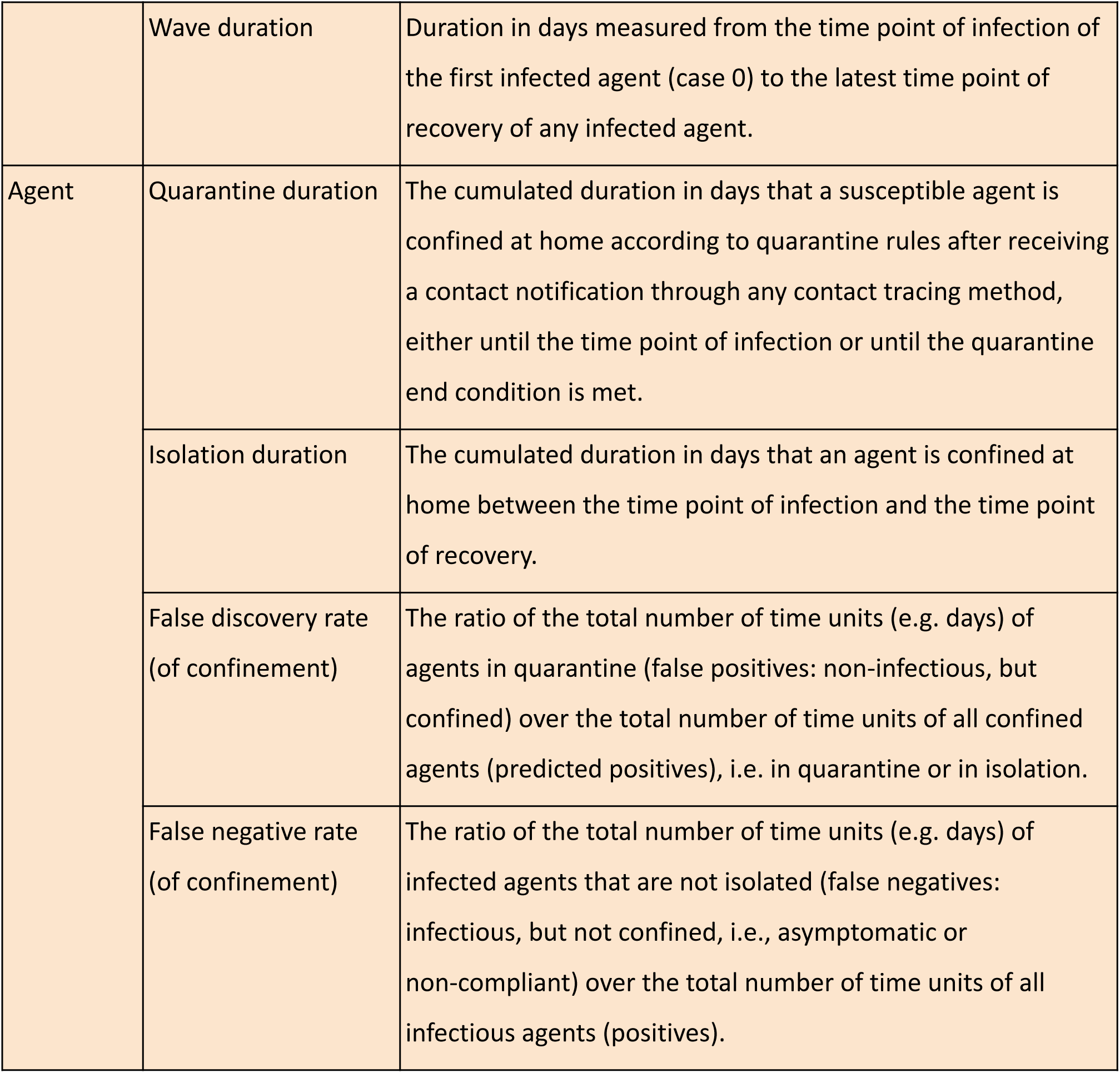

## Results

To analyse DCT effects, we present a prospective simulation-based analysis that builds on contacts emerging from an individual, behaviour-based modelling approach. We compare the simulation by its key components (population activities and behaviour, agent contacts, and pathogen propagation properties) against data from the literature. Subsequently, we performed a parametric search across the DCT adoption-adherence-compliance space and evaluated the parameters’ combined effect on key pandemic characteristics. Moreover, we show the effect of selected NPIs in combination with DCT in optimal and realistic behaviour scenarios.

Our simulation generates and tracks the physical positions of 1,000 agents in an appropriately sized virtual world, containing 500 apartments, 20 offices, 5 buses for public transport, as well as a hospital, a restaurant, and a bar (see Supplementary Fig. S1 [dct_analysis_supplement] and the Supplementary ODD protocol [odd_protocol] for further details). Simulations were performed with a time step of 5 minutes. Each pandemic simulation ran until there were no infectious agents in the population left (typically after 40 to 100 simulation days). Simulations were repeated 50 times for each parameter configuration; each simulation run had new agents and thus new home and office assignments, friends and acquaintances networks, and agent infectiousness profiles. At the beginning of each simulation run, five randomly selected agents were infected.

Using each agent’s position, we identify any encounter with another agent within a radius of 3m^30^ as a contact. If the contact occurred during the period of interest (e.g., previous 14 days for DCT and MCT) the agent of the contact becomes a contact person^31^. Contact duration refers to the time that any other agent stayed within the contact radius, counted in multiples of the simulation time step (5 min). Our default intervention is to isolate symptomatic agents after a positive test and release them after 14 days of isolation. In addition, we apply MCT and ICT in all simulations (for parameter configuration, see Methods section).

Our pathogen model captures important epidemiological concepts using agent states (susceptible, exposed, infected, immune, deceased), pathogene-related properties (viral load level and infectiousness level), as well as the disease and symptom states (see Fig. 1 for an overview). Based on established evidence that contact frequency, duration, and distance control infection^32^, we model an agent’s exposure mechanically. Specifically, an agent’s viral load level increases with time of exposure, when in contact with infectious agents, as a function of the contact’s distance and the contact person’s infectiousness level. Conversely, in the absence of contacts with infected agents, the viral load level of an agent progressively resets to zero, i.e., is washed out, according to a removal function. The transition from exposed to infected (i.e., self-sustaining viral state) is denoted by a critical threshold on the viral load level. An agent’s time-dependent infectiousness level controls disease progression (see Tab. 1 for disease states and characteristics). Details of the modelling can be found in the Methods section.

### Exploration of DCT effects

To explore DCT effects on a population during a pandemic, we simulated DCT parameter configurations for adoption, adherence, and compliance. A total of 38,400 simulation runs were performed for the DCT parameter exploration (8×8 DCT parameter grid points, 50 simulation runs per grid point, 4 pandemic characteristics, and 3 constant DCT parameter setpoints). We included MCT and ICT methods for all simulations, as they would coexist in a real scenario (MCT and ICT set to default parameters; see methodology section). Fig. 2 visualises simulation results for pandemic characteristics when sweeping DCT parameters. Results were averaged over 50 simulation runs per grid position and show an inhomogeneous optimisation space with various local optima. We derive that any path towards the global maxima across all DCT parameters will unlikely yield a monotonic improvement of the pandemic characteristics. With the complex parameter landscape, it becomes clear that the public perception of DCT effectiveness may be discouraged during its implementation, thus further inhibiting DCT adoption.

**Fig. 2:**
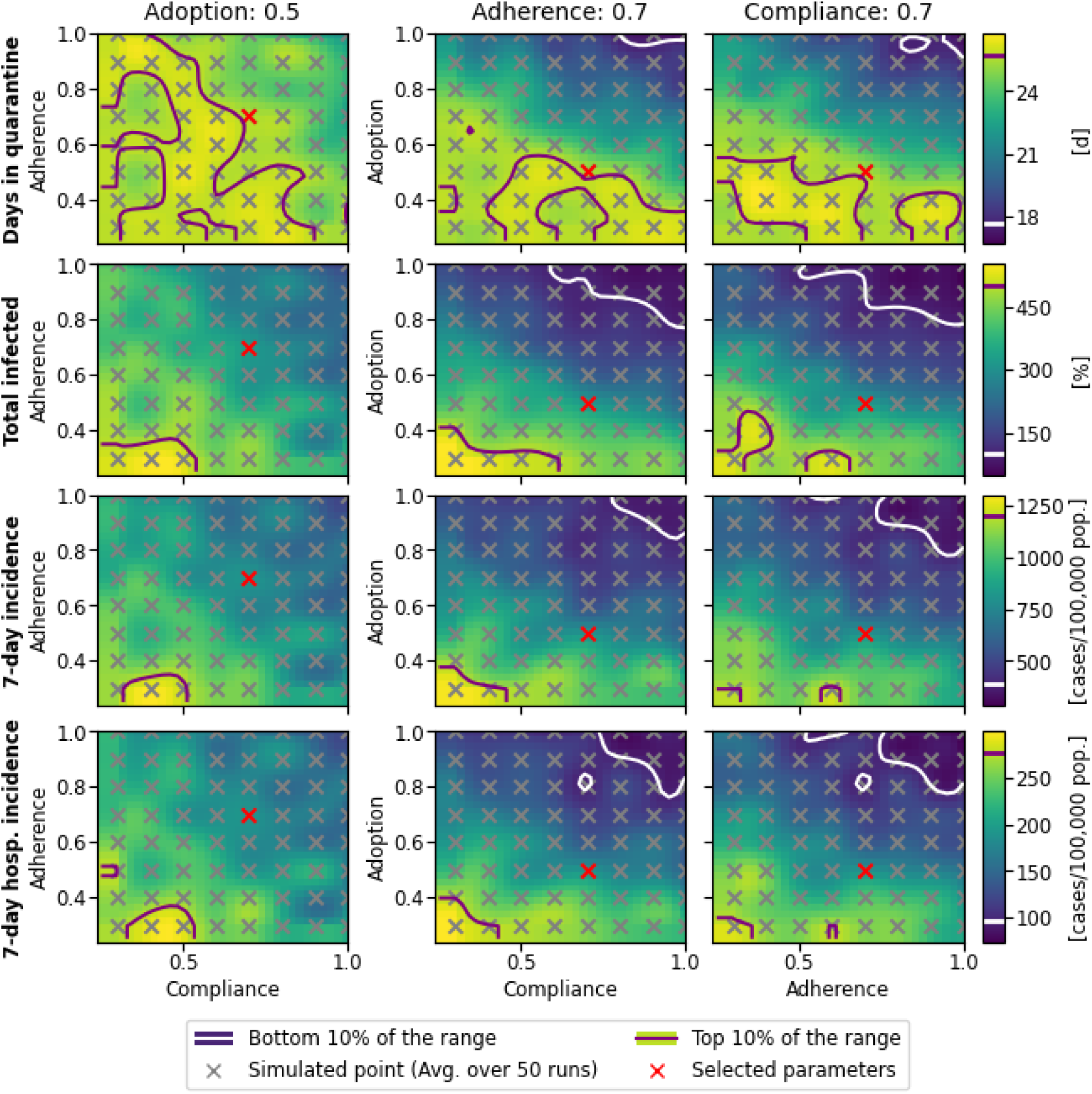
Exploration of DCT parameters: adoption, adherence, and compliance. Uniformly distributed parameter combinations were selected as simulated points. Results were averaged over 50 simulation runs per grid position. The first goal of a DCT implementation campaign should be to maximise adoption (activate DCT), followed by adherence (notify others), and compliance (follow recommendations). There is no monotonic path to improve pandemic characteristics, which presents a major challenge for any DCT implementation campaign.

In more detail, the parameter exploration suggests that DCT adoption has a key influence on the other DCT parameters (adherence and compliance). Thus, DCT adoption (i.e., user base) should be maximised first in a DCT implementation campaign. We subsequently evaluated pandemic characteristics at a DCT adoption of 50% to align with observations made during the COVID-19 pandemic^33^.

Based on the fluctuation of “Total infections” and “Days in quarantine” across the DCT parameter space, we conclude that DCT adherence and DCT compliance are similarly important. However, after maximising DCT adoption, our results indicate that prioritising adherence over compliance may lead to the lowest 10% region of the pandemic characteristics, even at an adoption far below the maximum. In more detail, the total infections and the 7-day incidence metrics approach a global minimum (low 10% boundary region), with compliance at 70%, adoption above 90%, and adherence as low as 50%. In contrast, fixing DCT adherence at 70% requires adoption above 70% to access the lowest 10% region.

### NPI interaction

We explored DCT and three NPIs (contact restriction or confinement relaxation) regarding their impact on pandemic characteristics. The NPIs modify the default NPI by inviting additional agents to quarantine (Quarantine Contact person Household - QCH), denying people access to specific public places (Closure of Bars and Restaurants - CBR), or changing the way normal activities are resumed (Require Negative Test to exit - RNT). Results were averaged over 50 runs per NPI combination. Thus, for 16 NPI combinations of realistic behaviour and 5 NPI combinations of DCT-optimal behaviour, a total of 1,050 individual pandemic simulation runs were performed.

Figure 3a shows pandemic characteristics in DCT-optimal behaviour scenarios (full DCT adoption, adherence, and compliance across the population). Two baseline simulations are shown: Baseline 1 does not apply any NPIs apart from MCT and ICT. Baseline 2 applies DCT in addition to MCT and ICT. The analysis shows that DCT, under DCT-optimal behaviour, can improve pandemic characteristics compared to Baseline 1. For example, without DCT, 60% of the population are infected, whereas with DCT, infections reduce to below 10% at half of the wave duration. Moreover, days in quarantine are less with DCT than without it. For the DCT-optimal behaviour scenario, additional NPIs do not show an additional effect on the pandemic characteristics, with the exception of RNT. RNT in addition to DCT can reduce the days in quarantine to about one third of the value obtained for Baseline 2. In summary, DCT has a profound effect on pandemic characteristics under DCT-optimal behaviour, which could be improved only in combination with RNT to reduce the days in quarantine thus mitigating the economic impact.

**Fig. 3:**
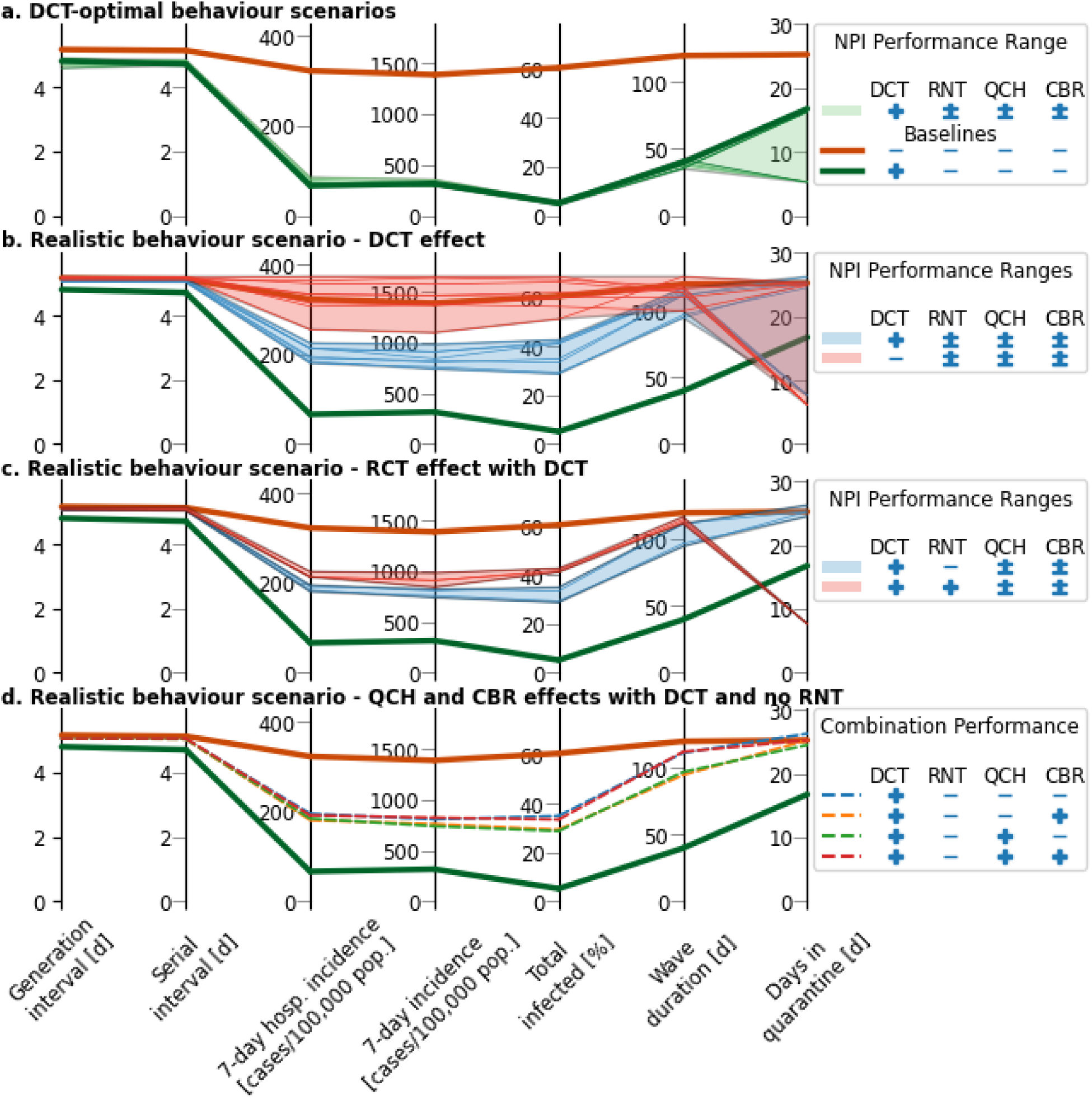
Exploration of DCT and NPI combinations regarding their impact on pandemic characteristics. Besides contact tracing, the following NPIs were considered: “Request negative test to exit” (RNT), “Quarantine Contact Person Household” (QCH), “Closure of Bars and Restaurants” (CBR). (a): DCT-optimal behaviour scenario (maximum for DCT adoption, adherence, and compliance). Baselines illustrate scenarios with and without DCT. Switching on and off RNT, QCH, and CBR creates a range of outcomes for pandemic characteristics. (b - d): Realistic behaviour scenario (DCT adoption: 50%, DCT adherence: 70%, DCT compliance: 70%). (b): DCT positively affects pandemic characteristics, even for realistic behaviour. (c): Exploration of RNT in addition to DCT. RNT does not influence pandemic characteristics. (d): Exploration of QCH and CBR in addition to DCT. Both QCH and CBR have a mutually exclusive effect on pandemic characteristics. (a - d): Shaded areas show the results range across active and inactive NPIs (denoted by “±”). All simulations include MCT and ICT. Incubation and clearance periods are not shown, as they are not affected by an NPI combination, i.e., the behaviour changes. Legend coding: (+) NPI active, (-) NPI inactive, (±) Both NPI active and inactive included. Results were averaged over 50 simulation runs per NPI combination. Simulation results for 7-day incidences are scaled to cases per 100,000 population. Definitions of pandemic characteristics can be found in Box 2.

Guided by estimations for the COVID-19 pandemic, we configured a realistic behaviour scenario (DCT adoption: 0.5^33^, DCT adherence: 0.7^34^, DCT compliance: 0.7^35,36^). Fig. 3b illustrates pandemic characteristic ranges for different NPIs when applied with or without DCT. “Request negative test to exit” (RNT) has a negative effect on pandemic characteristics as it sends agents back to the vulnerable pool. Our results confirm that RNT, without DCT, could perform worse than MCT alone. Fig. 3c illustrates worse pandemic characteristics when RNT is applied in addition to DCT. However, when DCT and RNT are combined, half of the quarantine days occur than without RNT.

DCT-optimal behaviour (Fig. 3a) lowers the serial and generation intervals to range at or below the median incubation time of ∼4.5 days. Hence, most interactions that lead to an infection occur at the end of the incubation period. In contrast, in the realistic behaviour scenarios, serial and generation intervals remain unaffected by DCT. Those 30% of non-compliant agents in our realistic behaviour scenarios with DCT applied, infected more people after their symptom onset compared to compliant agents. In particular, non-compliant agents increased the generation and serial intervals to 4.8 and 4.7 days, compared to 4.4 and 4.2 days in the DCT-optimal scenario. Due to the non-compliant agents, generation and serial intervals are similar to the baseline without DCT.

Different NPI combinations may yield similar results when applied together with DCT, as illustrated in Fig. 3d. DCT alone performs similarly to DCT with QCH and CBR. Furthermore, DCT and QCH perform similarly to DCT and CBR. The best pandemic characteristics are found for applying either QCH or CBR, but not simultaneously. Further interpretation is provided in the discussion section.

A side effect of DCT is that the median false discovery rate increases from 88.4% to up to 97% for DCT-optimal behaviour. When introducing DCT in realistic behaviour scenarios without RNT, the false discovery rate is ∼93%, depending on the combination of QCH and CBR. In contrast, the false discovery rate for realistic behaviour scenarios drops to ∼76% when using RNT in combination with DCT, and to ∼62% without DCT. Without DCT and RCT, combinations of QCH and CBR have a median false discovery rate of ∼88% (see Supplementary material [dct_analysis_supplement]).

In our simulation of realistic behaviour scenarios, the false negative rate was between 26% and 32%, and for DCT-optimal scenarios, the false negative rate varied between 24% and 36%. False discovery and false negative rates are shown in Tab. S3 for the DCT-optimal and in Tab. S4 for realistic behaviour.

We note that when applying RNT, the false negative rate was larger for the DCT-optimal than the realistic scenarios including DCT. By analysing the DCT-optimal scenario, we found that false negatives are generated by (1) unnotified asymptomatic agents, and (2) differences between our agent pathogen model and the standard DCT app behaviour. Due to the asymptomatic proportion of 40%, it is likely that infection chains of exclusively asymptomatic agents occur, which remain undetected by DCT. As RNT lets susceptible agents return to normal activities, they may get in contact with asymptomatic agents and hence get infected. In contrast, in a realistic scenario using DCT, non-compliant agents, while being infectious, continue to track their contacts. Thus, upon the contact between a non-compliant and an asymptomatic agent, DCT notifies the latter of an at-risk contact, as well as recommends testing and subsequent isolation of the asymptomatic agent.

Another reason for false negatives, even in the DCT-optimal scenario, is a discrepancy between our pathogen model and the simulated DCT app behaviour. According to our pathogen model, agents who accumulate sufficient viral load packages through multiple exposures of less than 15 minutes each will eventually get infected. However, our simulated DCT app follows the original WHO recommendations^37^, where infection is assumed after 15 minutes of continuous exposure, thus effectively failing to capture agents that got infected through multiple shorter exposures.

## Discussion

### Effectiveness of DCT and combinations with other NPIs

The focus of this work was to investigate the effects of DCT, both as a standalone measure and in combination with other NPIs. Our analysis demonstrates that DCT alone profoundly reduces pandemic impact in a DCT-optimal behaviour scenario with a 92% reduction of the total infections. In realistic behaviour scenarios, DCT still reduces total infections by one third compared to Baseline 1. Furthermore, our results show a substantial synergy between specific NPI combinations. In particular, combining DCT with CBR or QCH in realistic behaviour scenarios yields the best overall performance, i.e., reducing the total infections by 27%, from 394 for DCT only to 287 for DCT and QCH. Retrospective analyses^5^ on the importance of combining DCT with other NPIs corroborate our findings. However, even if individual NPIs have a positive effect on pandemic characteristics, their combination may not, as observed in our results when applying both QCH and CBR with DCT. As individuals spend more time in close proximity at one location, they may form a locally dense contact network and thus infect others. Here, QCH and CBR may have forced more agents to spend time at the home location. Further investigation with a behaviour-driven modelling approach could bring additional insights on NPI combination effects.

We found that DCT increases the false discovery rate, i.e., DCT leads to a higher proportion of individuals being quarantined. Conversely, DCT decreases the false negative rate, thus fewer infected individuals are left undetected and unisolated. Even more so, our results show that DCT confines more people per risk contact than any other NPI considered in the present analysis. Due to the comparably large false discovery rate of DCT, more of the asymptomatic individuals get notified, thus tested, and subsequently isolated, than without DCT.

Although DCT adoption plays an essential role in pandemic control^2^, our analysis confirms that maximum DCT adoption is unnecessary to reduce key pandemic characteristics. For example, an adoption of 50% in the realistic behaviour scenario yielded a reduction of the total infections by 42%. While DCT introduction campaigns should aim at improving adoption and adherence, pandemic characteristics do not react homogeneously (see Fig. 2), which may negatively influence public perception. Therefore, it is important to maintain ongoing public motivation strategies to offset any short-term setbacks. Additionally, it is not appropriate to judge the effectiveness of DCT solely based on short-term pandemic indicators, as they might give a misleading impression of DCT’s ineffectiveness in pandemic management.

In a typical ABM, the predefined social networks are characterised by their node degree, i.e., the contact person count per agent. Predefined networks are often static, i.e., not evolving over time. However, behaviour changes induced by NPIs change networks. The relationship between behaviour changes and network dynamics is often neither observed nor derived. Consequently, often unverified assumptions need to be made about how behaviour changes affect the networks. With a behaviour-driven ABM, we replace assumptions about how behaviour changes affect network dynamics by assumptions about pandemic-independent, verifiable behaviour patterns that can be altered by NPIs. In a behaviour-driven ABM, agent interactions create the network dynamics. Emergent networks allow us to evaluate metrics, including degree (i.e., contact person count) and strength (i.e., aggregated encounter duration across all contacts).

Our analysis of network metrics and pandemic characteristics revealed epidemiological implications. In particular, our network dynamics analysis (see Supplementary material [dct_analysis_supplement]) showed that infection time and 7-day incidence correlated with network strength rather than network degree. In addition, network strength reflects the change in contact preferences, if an NPI modifies individual behaviour. Consequently, the reported risk increase for household members due to isolation^38^ is not connected to the average degree of the family network. While the average degree of the family network did not change in our analysis, the average family network strength rose during isolations, which may explain the increased infection risk of household members during isolation periods.

DCT advances over MCT by removing recall issues associated with location, familiarity of contact persons, and time since last contact^39^. Furthermore, a DCT app could track short encounters and is not limited by workforce availability to collect and process contact information. While our settings for MCT did not elicit capacity limitations for the simulated population size, MCT did not overshadow DCT and the latter clearly improved pandemic characteristics. Based on our results, we conclude that DCT outperforms MCT risk assessment not only due to unrestricted capacity, but the reduction of contact processing time.

During the COVID-19 pandemic, the US CDC changed the initial contact tracing rule from 15 minutes of continuous exposure to a cumulative exposure duration of at least 15 minutes within 24 hours^40^. Our analysis of false negative cases confirms the CDC decision. More infectious pathogens may spread during shorter interactions. For example, at the emergence of the omicron variant, the US CDC further shortened the duration threshold to 10 minutes, due to the variant’s increased infectiousness^41^. At our current simulation time step of 5 minutes, the omicron variant spread could still be represented, but short exposure risk would be underestimated. The simulation time step could be reduced to represent short activities and their exposure risk. Besides the pathogen, DCT technology may affect the simulation time step. For instance, the Apple and Google Exposure Notification Framework^42^ implemented a dynamic delay between scans in the range of 2 to 5 minutes. For investigations on the sampling mechanism, a simulation time step of under 1 minute is justifiable. However, the simulation time step relates indirectly to the simulation time and processing costs. The simulation time step should be chosen carefully to balance simulation resolution with information gain.

The evaluation of guidelines for isolation and using rapid antigen tests identified that unnecessary quarantines produce an avoidable economic burden^43^. Therefore, days in quarantine can be considered a proxy for the economic burden of individuals for whom working from home is not an option. Our findings suggest that RNT is the only NPI that directly reduces the economic burden of quarantine. However, the quarantine reduction by RNT comes at the cost of increases in other pandemic indicators, including 7-day incidences, total infections, and wave duration (see Fig. 3c).

Mobility-restricting NPIs are disregarded in the public opinion. Thus, QCH might be more challenging to implement than CBR. However, our findings indicate that CBR and QCH offer limited differential impact on pandemic characteristics. Therefore, the economic benefit of keeping facilities operational, e.g., restaurants, might outweigh the costs associated with enforcing QCH and businesses losses.

### Modelling approach and metrics choice

Unlike traditional ABM models that rely on social network structure to describe the effect of NPIs, modelling an emergent network enabled us to analyse individual NPIs, including DCT, as well as NPI combinations by their direct effect on individual behaviour and compound effect on pandemic characteristics. To calibrate a behaviour-driven ABM, we performed several analyses of activity patterns, pathogen characteristics, contact patterns, and contact location (see Supplementary material [dct_analysis_supplement]). Moreover, we verified network dynamics for their plausibility regarding contact network shapes and the correlation between network metrics and pandemic characteristics (see Supplementary material [dct_analysis_supplement]).

It is recognised that human behaviour plays a critical role in pandemic development, from individual decisions to the level of policies, e.g., NPIs. Recent works by Wang et al.^44^ and Blanco et al.^45^ presented simulations of disease spread, including NPIs. While we share their objective to achieve realistic simulations of everyday behaviour within detailed small-scale cityscapes, including, e.g., offices and restaurants with typical environmental features and behaviours, our work significantly advances the field in several key areas. Our approach and analysis places a strong emphasis on epidemiological principles and metrics by proposing a novel viral load transfer model and key epidemiological metrics (see Box 2). Our approach combines epidemiologically informed scenarios (see Figure 2) with a behaviour-driven ABM. Blanco et al. asserted the potential of crowd simulation, in particular at the “atomic” level of individual agents where infection occurs. Our present analysis is explicit about emerging macro- and meso-level quantities (e.g., total infections and confinements stratified by notification type, see Supplementary video [supplemental_video_1]). In summary, our work breaks new ground in behaviour-driven modelling and simulation by integrating rigorous epidemiological principles and detailed ABM methodologies, thereby offering a robust and scientifically grounded approach for analysing disease spread and assessing public health interventions.

We introduced a novel pathogen model since no suitable representation of virus transfer existed. Our pathogen model represents distance and timing on continuous scales to implement the concepts of emitted and received viral load, including a viral load threshold and removal function that control the agent transition from exposed to infected. We separated the viral transmission from the contact generation mechanics. In total, ten pandemic characteristics (see Box 2) were used to validate the model against SARS-CoV-2 epidemiological data (see Supplementary material [dct_analysis_supplement]). With the new pathogen model we measured the effect of DCT and other NPI combinations.

While the selected pathogen characteristics are well established, e.g., the serial interval, there is no consensus on how to evaluate the combined effect of multiple NPIs. Here, we selected pandemic metrics that are generally recognised and used during the COVID-19 pandemic. Consequently, we did not emphasise metrics that apply to DCT only, e.g., turn-around times^10^. Several pandemic metrics (7-day incidence metrics, days in quarantine, and total infections) were derived due to their relevance for different stakeholders, including individuals, the health system, and the public.

A specific property of our approach is that pandemic metrics did not need to be specified upfront but emerged from the simulation. Our approach could be extended to design and test NPIs that have not been applied before. In particular, NPIs that depend on a person’s characteristics, e.g., household size and viral load profile, may be analysed with our approach and, more specifically, by reconfiguring the platform provided with this work. The NPIs would be comparably more challenging to describe in compartmental models, which assume that agents are interchangeable.

### Curse of complexity

Our current analysis was focused on western culture and daily repeated activity patterns in an urban setting, using office spaces, public transport, and bars and restaurants as a surrogate of informal inter-household interaction. Future studies could explore additional activity patterns influenced by culture, age, and work vs leisure days. Furthermore, other inter-household activities, including visits to other agents’ homes, could be added to provide additional infection pathways. Moreover, the pathogen model could be adjusted for virus variations. While our approach can be extended to more diverse conditions as described above, our current results illustrate the potential, limitations, and contributions of NPIs, including DCT, alone and in concert. Future work may broaden the scope and refine understanding while still managing complexity^46^. We argue that the flexibility of behaviour-driven ABM is two-sided. While our approach offers countless ways to explore uncharted conditions, it burdens the modeller with decisions on fidelity, thus potentially adding up structure without necessarily enhancing understanding and results.

### Prospects for NPI design

It is clear that modelling a virtual world will only approximate actual behaviour and facilities (see Fig. 1). For a behaviour-driven ABM, virtual world complexity, daily routines, location detail, activity diversity corresponding to routines and locations, as well as agent count, are main factors that affect simulation fidelity. Hence, a behaviour-driven ABM simulation could serve as a first exploration step when designing and orchestrating NPIs. Put differently, if an NPI is ineffective in the simulation, it is unlikely that the NPI will create benefits in reality. For example, current Bluetooth technology for indoor localisation provides imperfect measurements^13^. However, we showed that while assuming flawless wireless distance measurement and realistic agent behaviour, the effect of DCT on the pandemic is limited, with 35% of the population still getting infected. Therefore, the simulation results are a performance expectation boundary, where we would expect DCT implemented with current Bluetooth technology to perform worse than our idealised case.

For SARS-CoV-2, the original pathogen transmission characteristics were understood within six months after the initial outbreak^47^ and subsequent variants were characterised within weeks^48,49^. NPI effects on pandemic characteristics require additional time in extent of weeks to months to be determined in an ongoing pandemic^50^. However, quickly understanding the effects of NPIs is essential for policymakers to control pandemic development^51^. Moverover, the population may perceive an NPI and potential NPI adjustments as unnecessary grievances as they restrict personal freedom^52^. Furthermore, evaluating a single NPI’s effect on pandemic characteristics is infeasible as effects cannot be isolated in reality. Thus, behavioural simulation is a key approach to design and test NPIs regarding short-term and long-term effects and before implementing them in reality. Thus, our approach equips policymakers with a tool to understand an NPI’s effectiveness in terms of 7-day incidences and total infections, compared to collateral damage expressed in wave duration, days in quarantine, days in isolation, false discovery rate, and false negative rate. For example, policymakers can use the simulation to analyse the benefits of implementing RNT as a measure to reduce economic burden.

Since simulated agents are not required to match actual individuals, no privacy-critical data from the citizens are required to evaluate the NPI’s effectiveness. With behaviour-driven ABMs, no data from previous NPI implementations are necessary to create and simulate contact networks. Consequently, in a virtual world representation and assuming that the underlying viral transmission characteristics are known, the NPI analysis time depends on simulation capacity only.

**In conclusion**, behaviour-driven ABMs add important insight on DCT effectiveness as well as help to understand the impact of combining and fine tuning NPI combinations that maximise effectiveness, minimise collateral damage, and avoid more disruptive interventions, including proactive school closures or general lockdowns. Compared to predefined network ABMs, behaviour-driven ABMs are easier to adjust and to interrogate regarding specific environments, population routines, and virus properties, and provide additional transparency and explainability that helps to disentangle assumptions. For example, the complexity of behaviour patterns inside locations can be modified independently of the pathogen transmission mechanics, which helps identify risky behaviours, rather than assuming a priori that certain behaviours are risky. Future investigations may model further detail in the virtual worlds and consider additional metrics to measure social and economic impact. Based on the simulation results for a selected region of interest, policy design can balance pandemic impact with cultural differences across communities, thus preventing possibly discriminatory NPIs.

## Methods

ABMs serve as a pivotal tool for unravelling hypothetical outcomes in complex systems. Here, we investigated the specific and joint effect of DCT and other NPIs on pandemic scenarios, which are governed by contacts between people. A component-based model architecture facilitates our approach, which encapsulates individually validated, realistic properties. The architecture builds on three model layers: behaviour, pathogen, and NPI. To delve deeper into our model’s framework, encompassing its entities, parameters, and initial states, we guide readers to the ABM ODD (Overview, Design Concepts, Details) protocol^53,54^ [odd_protocol] and our Supplementary material [dct_analysis_supplement]. Specifics of the modelling and simulation platform can be found in our Python-based implementation, which is publicly accessible on GitHub [https://github.com/i2mb/dct-covid-bd-abm].

Behaviour drives agents to engage in various interactions within their social networks of family, friends, and acquaintances, and experience encounters outside these circles, thus providing additional risk of contracting or transmitting the virus. In greater detail, we simulate agents’ activity patterns—including the nature and duration of activities—considering variables such as daytime, the agent’s current location, infection state (viral load level, infectiousness level). Therefore, we conceptualise NPIs as modifiers of the individual behaviour that drives agent interaction, with the goal to reduce viral transmission. In other words, the NPI regime shapes the behaviour of infected agents, their contact persons, and the population as whole.

### Zeitgeber architecture

In our approach, agent behaviour is intricately shaped by various factors, including the cyclic nature of daily routines, environmental contexts, and intrinsic personal necessities. Thus, behaviour plays a pivotal role in guiding the agents as they navigate through the virtual world.

#### Zeitgeber

To realistically represent the daily life patterns of individuals, we model a Zeitgeber with three daily routines, each representing a segment of a typical day (refer to Fig. 1a). *Diurnal Activity* encapsulates a structured array of work-related activities, including commuting, office work, and work breaks. *Adaptive* is a daily routine with increased flexibility and adaptability, where agents may engage in evening activities, ranging from staying at home to social outings. *Nocturnal Recuperation* is dedicated to rest and recovery, essential for resetting the agents’ internal biological clocks. Each daily routine dictates agent location and sets the stage for their scheduled, reactive, and opportunistic activities^55^. For example, a scheduled activity (here equivalent to location-dependent default activity) is office work. A reactive activity is, e.g., to quarantine upon being requested by a contact tracing notification. Opportunistic activities are suggested by a situation or environment, e.g., to cook in a kitchen. To transfer between locations, agents schedule their commute, in particular, to transfer between work and home locations. For an exhaustive list of activities, along with their associated environments and types, we direct readers to the Table ODD1 in the Supplementary ODD protocol [odd_protocol]. The hierarchical structure of the Zeitgeber and the layers that synthesise agent activities are depicted in Fig. 5.

**Fig. 5:**
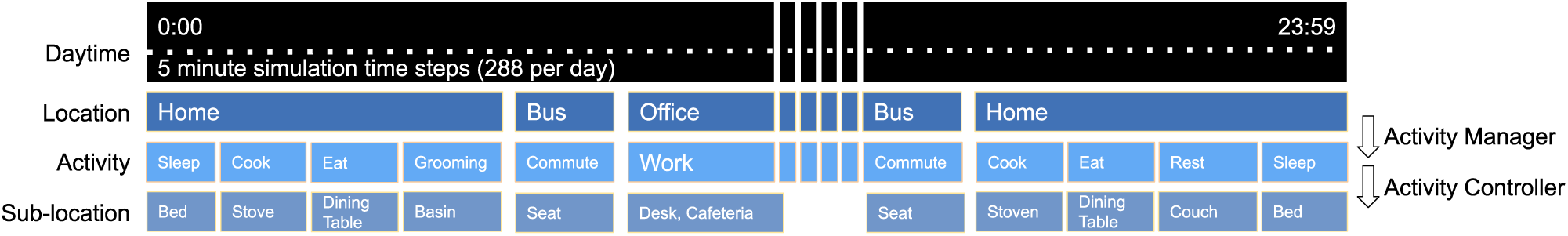
Example of a single-day activity trace illustrating the Zeitgeber architecture’s three contextual characteristics (daytime, location, activity). In our behaviour model, the Zeitgeber schedule determines location, the Activity Manager orchestrates activities represented by Activity Controllers, and Activity Controllers determine individual activities and their sub-location.

#### Activity Manager

The *Activity Manager* orchestrates activities at the agent’s current location based on daily routine and location determined by the Zeitgeber. When an agent arrives at a location, different effects influence their engagement in activities: (1) Intrinsic necessities represent an agent’s internal drive to undertake specific activities when feasible, e.g., to sleep when at home. (2) Available facilities, e.g., beds, stoves, etc., provide the basis for an agent’s interactions with their surroundings. For each location type, an activity list and required facilities were predefined. From an implementation perspective, the Activity Manager polls and prioritises location-based *Activity Controllers* that model an activity and its required facilities. By choice of design, NPIs were implemented as changes in schedule and location availability.

#### Activity Controllers

Location-specific *Activity Controllers* are used to determine activity patterns, i.e., onset and duration of an activity, its interruptibility by other *Activity Controllers*, as well as a delay (i.e., cool-down period) before repeating an activity. *Activity Controllers* encode knowledge about necessary objects and facilities as well as required motion patterns for the activity, thus can simulate intricate movements and interactions. For example, the *Activity Controller* for cooking requires kitchen furniture, and the controller for sleeping requires a bed.

A more elaborate example: When the Zeitgeber determines “home” as location, the *Activity Manager* polls all *Activity Controllers* of the home and may determine the agent to initiate cooking. The *Activity Manager* derives activity information from the corresponding *Activity Controller* for cooking, including moving the agent into the sub-location kitchen, positioning the agent appropriately in front of one or more kitchen appliances, etc.

For the *Activity Manager’s* orchestration of activities, sleep has the highest priority. For sleep at the agent’s home, sleep patterns are derived according to circadian distributions for sleep duration and sleep midpoint. If no other *Activity Controller* provides an activity during wake hours, the opportunistic activity (i.e., default activity) of the location is chosen. Thus, every agent receives an activity at all times during the simulation.

### Pathogen model

Our pathogen model captures relevant agent states (susceptible, exposed, infected, immune, deceased), pathogen-related properties (viral load level and infectiousness level), as well as the disease and symptom states. For details, see ‘Pathogen submode’ section in the [ODD protocol] and ‘Viral load threshold’ below. Table 1 offers a comprehensive overview of the key pathogen concepts and their relations throughout a typical infection. The model integrates empirical data to represent pathogen spreads and infection. Disease progression of infected agents is tracked in the simulation to assess NPI effectiveness.

#### Infectiousness level

*il*_*j*_(*t*) is the normalised infectiousness level of the *j*-th agent at time *t*. The infectious level determines the transmittable viral load to a susceptible or exposed agent (see also Eq. 3).

Technically, we use triangular functions to approximate *il*_*j*_(*t*). For each infectious agent *j*, an incubation period 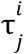, a clearance period 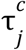 (see Box 2), and a peak infectiousness level 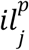 are sampled from the truncated normal distributions 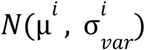, 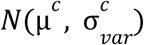, and 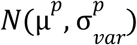, resulting in a unique triangular function per infectious agent *j* called the infectiousness level profile. The function for the *j*-th agent is given by:

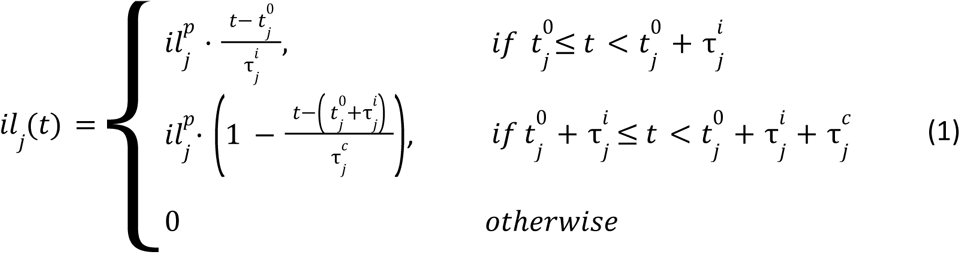

where 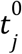 is the infection time for the *j*-th agent (see paragraph ‘Viral load threshold’ below).

The normalisation factor and distribution parameters are based on empirical data^56^ and provided in the Supplementary ODD protocol [odd_protocol]. We provide an exemplary visualisation of 50 sampled infectiousness level profiles in Fig. 6a; for visual convenience, we centred all infectiousness level profiles at the time point of symptom onset here.

**Fig. 6:**
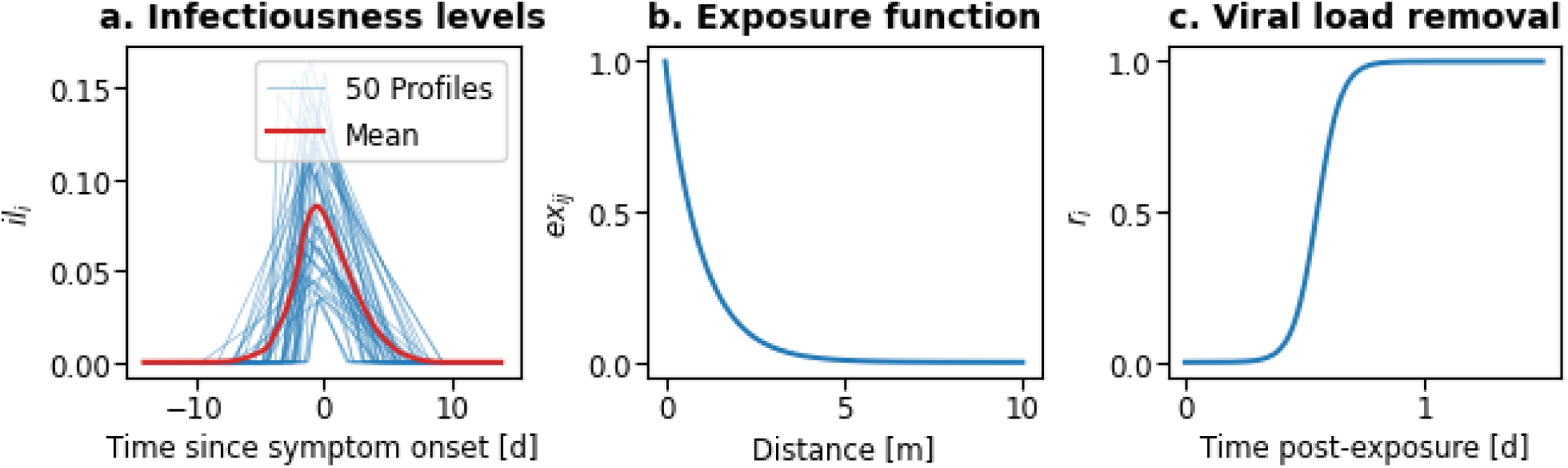
Key functions of the pathogen transmission model. (a) Infectiousness profiles of 50 exemplary agents, sampled from empirical data^56^. The profiles are relative to the agent’s symptom onset. (b) Viral exposure function to represent the distance-dependent decay of viral load transmitted from agent j to agent i (see Eq. 1). (c) Wash-out, i.e., time-dependent viral load removal, to represent an agent’s immune system reduction of viral load level.

#### Viral load level

The viral load level *vl*_*i*_ (*t*) of agent *i* denotes the pathogen quantity at time *t* and is pivotal in determining transitions between different agent states (susceptible, exposed, infected). Agents may accumulate viral load when in contact with infectious agents. Thus, our pathogen model aligns with evidence that frequent, enduring, and close exposures are more likely to lead to infection^32^.

The viral load package, transferred from infectious agent *j* to susceptible agent *i* is modelled according to the viral exposure function 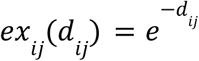, where *d_ij_* is the Euclidean distance between agents *i* and *j* (see Fig. 6b). A homogeneous radial spread pattern of viral load with exponential decay depending on the distance between agents is assumed (see also Fig. 1c). We calibrated the model with empirical data based on the large aerosol droplet propagation distance generated while speaking^30,57^. The approach approximates a more complex fluid dynamics simulation that would require virus particle tracking.

The viral load level (*vl*_*i*_) of a healthy agent *i* at simulation time *t* depends on the viral load level of the previous simulation time step *t* − 1, the newly received viral load packages, and the viral load removal according to 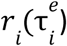, if there were no new encounters with infectious agents. The viral load transfer is determined by:

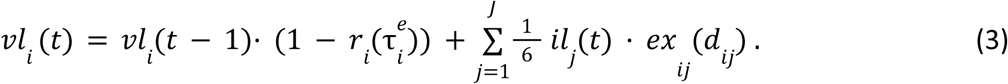

By design, the infectiousness level reflects the size of the viral population inside the host. By using the serial and generation intervals, we calibrated the viral load that is exhaled, to ⅙ of the infectiousness level at simulation time *t*. Please see Supplementary material for details.

#### Viral load removal

The wash-out mechanism is essential to represent the immune system’s function in reducing the previously accumulated viral load level. We model the process by introducing the viral load removal of agent *i* depending on the elapsed time since the last exposure (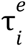, measured in days):

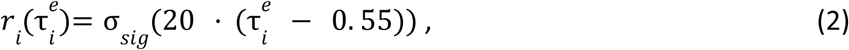

where σ_*sig*_ denotes the sigmoid function (see Fig. 6c). The viral load removal is based on a transmission case^58^, where short, multi-exposures, within 24 hours, led to the redefinition of a “close contact”. The function’s constants were calibrated by measuring the emergent serial and generation intervals.

#### Viral load threshold

A viral load threshold θ controls the transition from exposed to infected state. When an agent’s viral load level *vl_i_* (*t*) accumulates to the viral load threshold θ, the virus is considered self-sustained and no longer depends on accumulation and removal dynamics. When *vl_i_* (*t*) exceeds θ, infection time 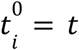 is logged for the newly infectious agent *i*. After recovering from the disease, agents are considered immune until the end of the simulation. Therefore, for any agent *i*, viral load level *vl_i_*(*t*) is undefined after 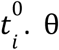 was set to 1 representing the average normalised viral load that an agent needs to transition from exposed to infected state. See the Supplementary material for details on the validation of the viral load threshold θ.

#### Diagnostic tests

Testing for the virus is triggered one day after symptom onset and conducted during business hours (8:00–16:30). Agents receive test results 24 hours after the test was conducted. Positive tests are communicated through contact tracing methods. Asymptomatic agents are not tested voluntarily, they only get tested under RNT after being isolated by contact tracing. Symptomatic agents will get tested on the next day after symptom onset. Agents with mild symptoms self-isolate, while severe cases are hospitalised.

#### Immunity and mortality

During the infected agent state, there is a small probability (2%) of mortality. Deceased agents are placed in the virtual cemetery. All recovered agents are immune for the remaining simulation time.

### NPI Models

NPIs are conceptualised as a spectrum of actions individuals (can) take, i.e., to prevent infection, including using a DCT app (see Box 1 for an introduction). Each simulated agent has a behaviour profile representing the probability of undertaking specific NPI actions. NPIs are integrated as modules into the simulation framework, where they are treated as extensions of the Zeitgeber.

#### Contact tracing models

DCT has been introduced as part of the NPI portfolio in many regions at the onset of the COVID-19 pandemic. In particular, DCT is hypothesised to provide an alternative that overcomes the limitations of MCT^2^. In the present work, we modelled three different contact tracing strategies: DCT, MCT, and ICT (see Box 1). In our implementation, all contact tracing strategies adhere to a uniform process flow: (1) categorise contacts, (2) construct contact networks, (3) acquire positive diagnostic tests, (4) consider to inform contact persons, and (5) consider to comply with recommendations. Subsequently, we provide more conceptual detail of the process flow implementation.

#### Contact categories

Contacts are categorised at the beginning of the simulation, depending on the contact tracing strategy. For DCT, all contacts are considered equal. MCT splits contacts by location into office contacts and home contacts. For ICT, we considered family contacts (with agents from the same household), friends contacts, and acquaintances contacts, where friends and acquaintances are agents selected from the same office.

In our behaviour-driven modelling, the shape and strength of the contact networks are an emerging property (see Supplementary [dct_analysis_supplement]: Network Dynamics and subsequent sections). We analyse contact types by categorising the pairwise relationships between all agents. Initially, the number of agent pairs selected for the friends category is 40% of the office size. Accordingly, in a 20-person office environment, the friends category is constructed by randomly selecting eight agents. To construct the acquaintances’ contacts, 50% of the office-sharing agent pairs that do not belong to the friends category were selected. During simulation time, ICT considers agents as an unknown contact person, if the corresponding agent does not fall within the friends and acquaintance contact persons.

#### Contact network construction

To construct contact networks, we consider a 3 m radius of contact candidates. For DCT, the minimum continuous contact duration was set to two simulation time steps, i.e., 10 min, before a contact within the radius is registered. For MCT and ICT we do not apply a contact duration constraint. Thus, we support that an agent might register a contact person although it was a brief encounter only.

#### Logging duration

Regardless of the contact tracing strategy employed, we log all contacts for a period of two simulation weeks, i.e., τ_*log*_ = 14 *days*. Contacts older than τ_*log*_ are considered “forgotten” and are no longer factored into the contact tracing process. The approach aligns with human memorisation and record-keeping limitations in real-world contact tracing scenarios.

#### Recall bias

Specifically for ICT and MCT, we model a recall probability to represent the temporal effects in recall performance for contacts. As a contact between agent *a* and *b* ages, the likelihood of recall declines. On the contrary, more recent contacts are more easily remembered; a phenomenon well-documented in cognitive psychology^39^. Eq. 3 expresses the temporal encounter recall *P*_*tr*_(*t*, (*a*, *b*)) between agents *a* and *b* as a function of the time since the last contact *t*_*lc*_, the cumulative contact duration τ (*a*, *b*), and the logging duration τ_*log*._

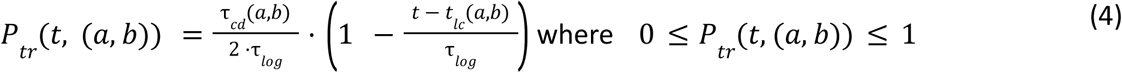

If *t* – *t*_*lc*_ > τ_*log*_ the contact is forgotten and τ_*tr*_ (*a*, *b*) = 0. The term 1/ (2 · τ_*log*_) downweights the cumulative contact duration τ_*cd*_ compared to the elapsed time since the last contact *t*_*lc*_.

The contextual encounter recall *P*_*cr*_(*c*, (*a*, *b*)) represents the situational probability to recollect the contact between a and b. In MCT, *c* represents the location of the contact (e.g., home, office, public place). In ICT, *c* represents the contact category (e.g., with family, friend, acquaintance). Contacts with unknown contact persons cannot get recollected in ICT. Contextual encounter recall *P*_*cr*_ was predefined per context *c* and can be found in the Supplementary ODD protocol [odd_protocol]. Combining both, the contextual and temporal encounter recall yields the total recall bias *P*_*rb*_ (see Eq. 5).

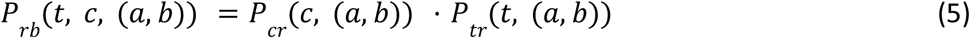

#### Adherence

A key success factor of contact tracing is whether individuals inform others of positive test results. Adherence was implemented as the individual’s probability of reporting a positive test result within their contact networks. In our approach, the contact tracing strategy does not affect an individual’s decision to report the test result, i.e., an individual who decides to adhere will inform all contact persons via DCT, MCT, and ICT. Consequently, non-adherence means that no contact tracing strategy could inform an individual of a high-risk contact.

#### Contact tracing capacity

For MCT, we model processing capacity limitations by constraining the message dissemination to an agent’s contact persons. In more detail, we simulate that agents report their infection state and a list of recalled contact persons to health authorities. Agent reports are held for three days to simulate MCT processing delay. Subsequently, contact person notifications are extracted from the reports and queued according to the reported time of contact. The queue is processed one notification per simulation time step. We simulated notification failures by applying an 80% success rate to notifying the contact person. Failed notifications are retried two times at most. Notifications remaining in the system that are older than five days are dropped from the queue. The capacity of delivering up to 288 notifications per day is equivalent to that of a typical German local health authority (Gesundheitsamt), which - at times of high incidence during the COVID-19 pandemic - was supported by one or two containment scouts for a population of 100,000 individuals^59^. In our simulations, the MCT capacity settings were applied to just 1,000 agents.

#### Compliance with NPI recommendations

Compliance varies across contact tracing strategies and is governed by compliance parameterisation. DCT compliance (see Box 1) was varied to contrast DCT-optimal and realistic behaviour (see Fig. 3). For MCT, we assume a uniform compliance rate of 0.95 across all agents, thus representing a general willingness to cooperate with health interventions. While MCT compliance reflects the perceived societal responsibility and trust in governmental organisations, ICT compliance varies across contact categories, acknowledging the differences in personal relationships. In particular, ICT compliance was 0.99 in case of family contacts, 0.9 for friends contacts, and 0.5 for acquaintances contacts, indicating varying levels of trust and social obligation across the contact categories.

### Intervention models

The objective of contact tracing technologies is to identify and isolate infectious individuals and quarantine their contact persons in order to curtail virus spread by reducing contacts between individuals. In our simulation, we conceptualised interventions as modifications to an agent’s usual behaviour. When an agent tests positive, they enter isolation and may notify agents in their contact network, thus triggering contact persons to quarantine. During confinement, i.e., isolation or quarantine, scheduling models that dictate an agent’s location changes are suspended. Agents resume their regular routines based on specific quarantine and isolation exit protocols.

#### RNT

Under the RNT intervention, confined agents are required to obtain a negative test result before returning to normal activities prior to the default 14-day delay. We simulate a waiting period of 1.5 days between the test being performed and the agent receiving the test result. Without RNT, the standard recommendation for confined agents is to wait 14 days before resuming normal activities. We implemented RNT by controlling the agents’ testing frequency.

#### QCH

During the COVID-19 pandemic, lockdowns and the emphasis on isolating and quarantining at home led to households becoming primary infection locations^38,60^. Our QCH intervention explores the potential impact of quarantining entire households when a member is isolated or quarantined. Thus, QCH aims to contain the virus spread within a single household, preventing further transmission to other households. QCH is implemented by using the Zeitgeber to stop allocating location changes for all agents in the household.

#### CBR

Closure of social facilities, e.g., bars and restaurants were prevalent during the pandemic^52,61,62^. To implement CBR, we disabled the scheduling model that facilitates agents’ night-out activities. We focused on a static scenario, i.e., the status of gastronomy services remains constant throughout the simulation run, thus reflecting the continuous closure of these establishments during certain periods of the pandemic.

#### Quarantine rules

Agents go into quarantine after receiving a contact notification through any contact tracing strategy and QCH, if applicable. The agent exits quarantine if one of the following conditions are met: (1) after the regular 14 days quarantine period has passed, (2) after receiving a negative test if RNT is used, (3) after all household members test negative, if QCH and RNT are applied together, and (4) by becoming infected, in which case, the agent is considered isolated and no longer quarantined. Recovered agents do not quarantine or comply with QCH after receiving a notification.

#### Isolation procedure

Symptomatic agents wait 1.5 days after symptom onset to get tested. Tests can be performed only during the test centre opening hours between 8:00 and 16:30. Agents receive test results after 24 hours. After receiving a positive test result, the agent may go into isolation.

## Supporting information

ODD protocol for the I2MB simulator

Supplemental material including validation methodology

## Data Availability

The code and procedure to regenerate the data are available at https://github.com/i2mb/dct-covid-bd-abm

## Acknowledgements

The authors would like to thank all the organisers and participants of the research community meetings on epidemiological modelling hosted by the University of Münster (Germany) from 2020 to 2021 for their comments and suggestions on how to elaborate our behaviour-driven ABM approach.

## Author contributions

L.I.L.G. developed the methodology, implemented the simulation, provided the visualisations, and ran the experiments. G.Kö. generated the ODD protocol and the overview Figure 1. G.Ki. advised regarding CoronaWarnApp experience and epidemiological modelling. J.B. advised regarding the CoronaWarnApp and epidemiological principles. O.A. developed the methodology and advised regarding behaviour and activity patterns. All authors advised and discussed the methodology, analysed the results, and contributed to the writing of the paper.

