## Supplementary material for "Impact of Digital Contact Tracing on Pandemic Control Analysed with Behaviour-driven Agent-based Modelling": ODD protocol for the I2MB simulator

### Overview, Design concepts and Details (ODD) protocol for the paper "Impact of Digital Contact Tracing on Pandemic Control Analysed with Behaviour-driven Agent-based Modelling"

Luis Ignacio Lopera González [1], Göran Köber [2], Göran Kirchner [3], Justus Benzler [3], Oliver Amft [2,4]
[1] Friedrich Alexander Universität Erlangen Nürnberg
[2] University of Freiburg
[3] Robert Koch Institut
[4] Hahn Schickard

Overview 1
Purpose and patterns 1
Entities, State variables, and Scales 1
Process overview and scheduling 2
Design concepts 2
Basic principles 2
Emergence 2
Adaptation 2
Objectives 2
Learning 3
Prediction 3
Sensing 3
Interaction 3
Stochasticity 3
Collectives 3
Observations 3
Details 3
Initialization 3
Input data 4
Submodels 4
1. Zeitgeber architecture 4
2. Pathogen submodel 6
3. NPI Submodel 7
State variables 7
Agent 7
**References 8**

### Overview

#### Purpose and patterns

This behaviour-driven agent-based model (ABM) is to simulate the dynamics of COVID-19 transmission in a population, considering the interactions between individual behaviours, pathogen transmission, environmental factors, and non-pharmaceutical interventions (NPI, see Box 1 in the paper). Specifically, the purpose of our behaviour-driven ABM is to simulate the dynamics of COVID-19 transmission depending on contact tracing strategies and to contrast them with contact restriction strategies and investigate their effects on selected pandemic characteristics (see Box 2). We discuss the results of our calibration analyses in the supplement.

#### Entities, State variables, and Scales

##### Entities

- Individuals/Agents in the population.
- Environments
  - Home: 500 homes, place to rest and sleep
  - Bus: 5 buses, commuting
  - Cars: commuting
  - Office: 50 offices, place to work and meet colleagues
  - Bar: meet with friends, encounter strangers
  - Restaurant: meet with friends, encounter strangers
  - Hospital: treat agents with critical corona symptoms
  - Cemetery: bury the dead
- Weighted agent networks
  - Family: household contacts
  - Friends: close out-of-household contacts
  - Acquaintances: recognizable out-of-household contacts
  - Stranger: none of the above

##### State variables

- Health status (dynamic; susceptible, exposed, infected, immune, deceased).
- Viral load (dynamic, float).
- Infectious level (dynamic, float): If infected, the disease follows a triangular function.
- Symptom levels (static; asymptomatic, presymptomatic, mild, strong, critical).
- Behaviour parameters (dynamic, current and past)
- Environmental factors (dynamic, current location)

##### Scales

- Spatial units (500 homes, 50 offices, 5 buses with 35 seats, cemetery, hospital).
- Time steps (5 minutes)
- Simulation ends with end of pandemic, i.e., when no exposed or infectious agents are left.

#### Process overview and scheduling

We designed a modular simulation engine (I2MB) that relies on three main model components: (1) simulated region, (2) behaviour and activity models, and (3) pathogen model. The component models are implemented in a modular fashion, i.e. they work independent of each other. Per simulation time step, models get sequentially applied on a shared data structure that represents the world state. For the present investigation we chose a five-minute simulation time step that provides sufficient resolution to represent social contacts and viral load transfer according to established literature[1–3].

1. Initialise the population with specified characteristics (locations, infectiousness profile, family and friend networks, office networks, home and office assignments, etc.).
2. Observe simulation time.
3. Update environmental states (closing times, available seats in e.g. a restaurant, etc.).
4. Check the schedule for location changes (home, office, bars and restaurants).
5. Simulate individual behaviours including motion and activities (scheduled, reactive, opportunistic).
6. Update pathogen infectiousness level and viral load level (exposure and removal).
7. Update NPI history.
8. Repeat steps 2-8 for each time step.

### Design concepts

#### Basic principles

- Individuals exhibit diverse behaviours influenced by the Circadian Activity Architecture, current location, and personal characteristics (health status, activity history, NPI).
- Pathogen transmission is modelled through interactions between infected and susceptible individuals in a time and space sensitive manner.
- Environments shape the behaviour of individuals.
- NPIs modify individual behaviours and transmission dynamics.

#### Emergence

- Emergent properties of interest include all population-level pandemic characteristics but clearance and incubation period (see Box 2 [dct_analysis]) and the impact of interventions on them.

#### Adaptation

- Individuals adapt their behaviours based on the regime of NPIs, their health status, their current environment, the day time, and history of behaviour.

#### Objectives

The model includes no objective-seeking behaviour.

#### Learning

The model includes no agent-sided learning.

#### Prediction

The model includes no agent-sided prediction.

#### Sensing

- Agents are aware of their current environment and the day-time and align their activities accordingly.

#### Interaction

- Direct interactions between agents appear in all environments but the hospital, car, and cemetery. Agents meet agents outside their households in bars, restaurants, their workplace, and public transportation. Agents meet their friends in bars and restaurants and family members at home.
- There are no indirect interactions between the agents.
- Communication between the agents about the infectious state happens depending on the NPI(s).

#### Stochasticity

- We repeated the runs 50 times with the same starting conditions and report the averages of selected pandemic characteristics (see Box 2).

#### Collectives

- Families: Agents of the same household.
- Friends: 40% of the office size (as an emerging property).
- Acquaintances: 50% of the office size which has not been selected as friend (as an emerging property).

#### Observations

- All population-level pandemic characteristics (see Box 2) and the impact of interventions are logged during the simulation.

### Details

#### Initialization

- Initial population n = 1000 agents (as default).
- n * .5 = 500 homes (distributed according p = [0.42, 0.33, 0.12, 0.09, 0.03] for 1-, 2-, 3-, 4-, 5-person households[4])
- n // 20 = 50 offices[5]

- n * .005 = 5 buses with 35 seats each (accommodating 12% of the population, the rest commutes by car[6])
- Initially infected: 5 (symptomatic). More detailed, the virus was introduced into the simulated population by randomly selecting an agent and setting its status to symptomatic, sampling a disease duration from the distribution, and selecting the disease outcome: immune or deceased. The first five agents were infected after the first step of the simulation and subsequent infections were driven by agent interactions.
- Agents are randomly assigned to households, workplaces, educational institutions.
- Simulation runs per analysis: 50

We generated our region using the desired population size in relation to the basic infrastructure ratios from Germany[7–9] and randomly distributed it into homes and offices per run. We further defined two hierarchy levels of location. We refer to location as the higher hierarchy level, e.g., home, office, or restaurant, and to sublocations as the spaces or rooms within a location. When synthesising activity traces, location inertia needs to be considered, while sublocation inertia is disregarded. Location inertia refers to how likely an agent is to change locations. For example, while we would avoid going back and forth between home and the office, we would have no problem switching rooms at home. To synthesise activity traces, we exploit the inertial difference between locations and sublocations, by scheduling location changes, while letting an activity manager choose between the available activities at the location to determine the agent's sublocation and its relative position.

#### Input data

- There are no external inputs to the model.

#### Submodels

The submodels are introduced in the methods section. Here we provide further details on behaviour and pathogen submodels and describe the virtual world.

##### 1. Zeitgeber architecture

Our Zeitgeber Architecture can also be seen as a time and location-aware behaviour model. The agents in our simulation engage in activities considering daytime, surroundings factors, and personal needs; these factors significantly influence their navigation in the virtual world. To mimic a typical day, we establish a Diurnal Activity (9 am – 5 pm) and an Adaptive Period (5 pm – 2 am or alternatively sleep onset). During the Diurnal Activity, unless isolated or under quarantine, agents have scheduled activities (commutes) to their full- or part-time workplaces. Nocturnal Recuperation is dedicated to rest and recovery (as depicted in Figure 1 of the article).
At their workplace, agents immediately engage in scheduled activities that can be flexibly interrupted by opportunistic activities (e.g., a break) to accommodate individual needs. Similarly, other environments have different schedules (e.g., in a restaurant agents eat, in the apartment agents rest) and opportunistic activities. Put differently, both scheduled and opportunistic activities are highly responsive to the environment. Reactive activities restrict the scope of available activities as a response to the NPIs, e.g., quarantine. For a comprehensive list of activities and their associated environments and types, see Tab ODD1.

| Activity Name | Location | Primary Type |
|---|---|---|
| Sleep | At home | S |
| CommuteBus | Bus | S |
| CommuteCar | Car | S |
| Work | Office | S |
| CoffeeBreak | Office | O |
| Eat | At home | O |
| EatAtBar | Bar | O |
| EatAtRestaurant | Restaurant | O |
| Rest | At home | O |
| Toilet | At home | O |
| Grooming | At home | O |
| Shower | At home | O |
| KitchenWork | At home | O |
| Quarantine | At home | R |
| Isolation | At home | R |
| Note: O = opportunistic, S = scheduled, R = reactive | | |

*Table ODD1: List of available agent activities and their related locations.*

Work Schedule:

- Schedule: 8:00@Office - 5:00@Home
- Repeats: Daily
- Applies to population percentage: HO - 100%, EX - 88%

Public Transport Schedule:

- Schedule: 7:15@Bus - Destination Office, 16:30@Bus - Destination Home
- Ride duration: Sampled from a uniform discrete distribution with values 15, 30, 45, and 60 minutes
- Applies to population percentage: 12%[6]
- Repeats: Daily
- Module mechanics: At departure times (@bus), commuting agents are randomly assigned a bus and a ride duration. Once the ride duration time has elapsed, agents are moved to their destination.

**Global motion:**

- Step size: 0.2m
- Mechanics: random motion with attractor fields.

**Night out:**

- Group location: Home or office
- Venues: Bar or restaurant
- Arrival: Sampled from a normal distribution with μ: 19:00 and σ: 1.5 hours
- Duration: Sampled from a normal distribution with μ: 4 hours and σ: 2.5 hours
- Opening hours: 17:00

- Closing hours: 02:00 next day
- Min capacity: 0
- Max capacity: the smaller of total available sitting capacity or 70% of the population
- Repeats: Daily
- Number of agents going out: uniformly sampled from the range min capacity max capacity
- Module mechanics: Agents are selected to go out at 6:00 pm, and the agents are grouped with other agents going out from the group location selected at random. The groups are assigned the same arrival time, and each agent is assigned an individual departure time. If there is not sufficient room to accommodate the entire group in the venue, their entrance is delayed by an hour. In the meantime, agents remain at home. This module enforces the closing and opening hours.

#### 2. Pathogen submodel

**Agent states**

**Susceptible:**

- Per default, all but 5 (symptomatic, see Initialization) initialised agents are susceptible to the virus.

**Exposed:**

- Agent that currently has accumulated viral load but is below the viral load threshold.

**Infected:**

- The agent acquired a viral load level greater than the viral load level threshold, and the virus is considered self-sustained in the agent.
- Agent contagion level is determined by the infectiousness level function.
- State duration: Sampled from a normal distribution with μ: 5.1 days and σ: 0.7 days[10].

**Immune, deceased:**

- After the disease has run its course.
- State duration: The rest of the simulation.
- Ratio: 2% death rate[11].

**Symptomatic, asymptomatic:**

- Agents are considered infectious.
- They can spread the virus to susceptible agents.
- State duration: Sampled from a normal distribution with μ: 15 days and σ: 4 days[12].
- Includes 2 days of the pre-symptomatic stage [13,14].
- Ratio: 40% asymptomatic [15,16].

**Symptom levels for symptomatic agents:**

- Assigned at the moment of exposure.

- Constant throughout the course of the disease.
- Sampled from a discrete distribution – asymptomatic: 40%, mild: 40%, strong: 13.8%, and critical: 6.2% [17].

**Additional parameters:**

- ICU beds: 3% of the population.
- ICU saturation death rate: 5%.

#### 3. NPI Submodel

We set the recall probability for home contacts $cm_{MCT}(a, b, c = home) = 0.98$, acknowledging the frequent and close nature of home interactions. We set $cm_{MCT}(a, b, c = office) = 0.5$ to express the more frequent and sporadic nature of typical office contacts. Public place contacts, often facilitated by gastronomy services collecting visitor information, were set to a recall probability of $cm_{MCT}(a, b, c = public) = 0.3$. The setting reflects the contribution of public establishments in aiding manual contact tracing efforts, e.g., bar and restaurant personnel providing the otherwise-unknown contact information. ICT provides further differentiation with recall probability as follows: $cm_{ICT}(a, b, c = family) = 0.99$, $cm_{ICT}(a, b, c = friends) = 0.8$, and $cm_{ICT}(a, b, c = acquaintances) = 0.4$. Thus, ICT recall probabilities express differences in varying degrees of interaction intensity and frequency among different social circles.

### State variables

#### Agent

| Variable name | Variable type and units | Meaning |
|---|---|---|
| activity | object, dynamic | one of the activities from the list above |
| AgentState (UserState) | categorical, dynamic | Levels for the agent states: susceptible = 0, immune = 1, deceased = 2, exposed = 3, infected = 4, infectious = 5 |
| SymptomLevels | categorical, dynamic | not_sick = -1 (not infected), no_symptoms = 0 (asymptomatic), recovering = 1, mild = 2, strong = 3, critical = 4 |
| infectiousness_level | float, dynamic | Viral load as a function of exposure time, exposure distance, and removal |
| isolated | bool, dynamic | Agent confinement true/false |
| location | categorical, dynamic | Office/schools, rooms (apartment components), bars, restaurants, hospital, cemetery, car, bus |
| position | xy-coordinates | Location-dependent coordinates of the agent |
