## Supplemental material including validation methodology for "Impact of Digital Contact Tracing on Pandemic Control Analysed with Behaviour-driven Agent-based Modelling"

Luis Ignacio Lopera González [1], Göran Köber [2], Göran Kirchner [3], Justus Benzler [3], Oliver Amft [2,4]
[1] Friedrich Alexander Universität Erlangen Nürnberg
[2] University of Freiburg
[3] Robert Koch Institut
[4] Hahn Schickard

#### Table of content

**Introduction** 1
**Virtual world infrastructure** 2
**Validation and calibration** 3
  **Validation of daily activity patterns** 3
  **Validation of contact patterns** 5
    **Overall contact patterns** 6
    **Location-based contact patterns** 7
    **Conclusion on contact pattern validation** 9
  **Validation and calibration of pathogen characteristics** 9
**Network dynamics** 10
  **Contact network shapes** 11
  **Correlation between network metrics and pandemic characteristics** 14
  **Analysis of daily contacts, degree, and strength** 19
**False discovery rate and false negative rate** 23
**DCT-optimal behaviour** 25
**Realistic behaviour** 26
**References** 27

#### Introduction

This supplemental material presents a comprehensive investigation on the validation and calibration of our behaviour-driven agent-based simulation engine, termed individual-to-mass behaviour (I2MB). We start by showing how the virtual world is constructed and detailed the virtual world's infrastructure, followed by the validation of daily activity patterns, contact patterns, and the pathogen model. Subsequently, we investigate agent interaction behaviour and its effects on selected emerging pandemic characteristics using network dynamic analysis. Finally, we present tables of false discovery rate and false negative rate as referenced in the main article.

The I2MB simulator was implemented in Python and follows a modular design. The main article presents the abstractions and the theory behind the I2MB implementation. The technical implementation details for the I2MB engine and each module can be found in the accompanying Overview, Design Concepts & Details (ODD) protocol and GitHub repository.

#### Virtual world infrastructure

Given the characteristics of the COVID-19 virus and the comparably low risk of outdoor contagion, our simulations focused on indoor locations. For simplicity, the hospital and cemetery do not have internal partitions since the interaction between agents at these locations does not contribute to virus transmission and they can be disregarded in contact statistics.

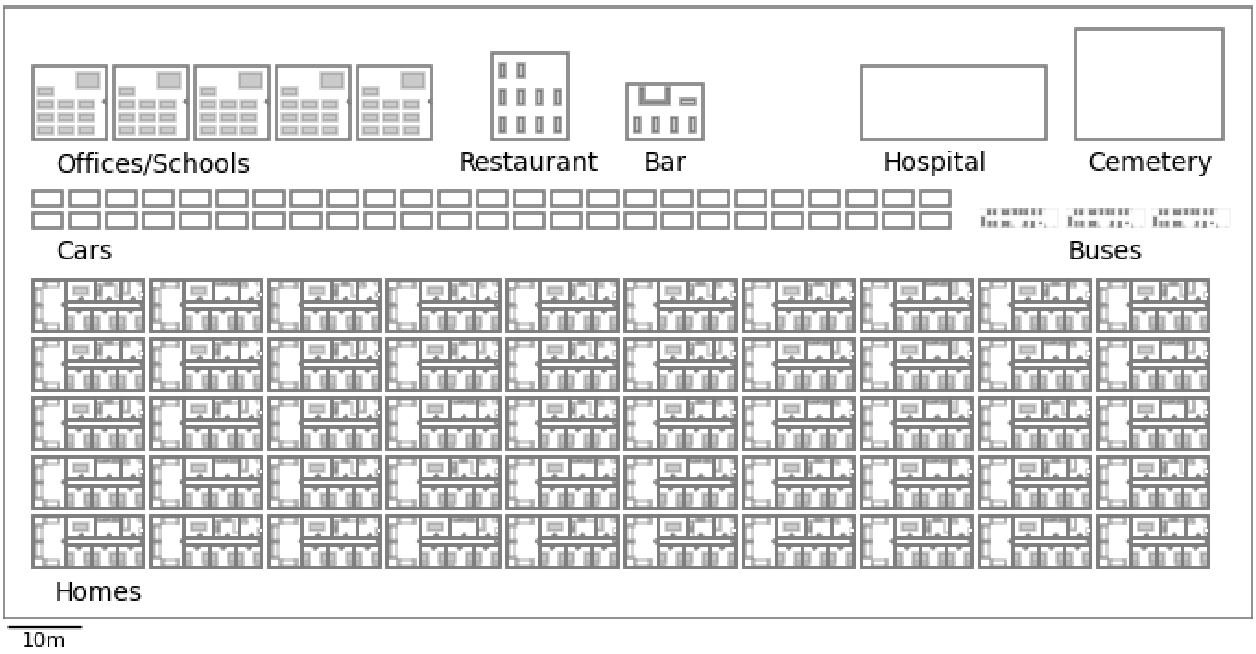


*Fig. S1:* Illustration of the virtual world infrastructure scaled for 100 agents. Life outside of the modelled spaces, e.g. homes, cars, etc. was not considered.

The virtual world depicted in Fig. S1 was scaled proportional to the population (see ODD protocol for initialisations). Agents were assigned homes such that the number of agents per home distribution matched the 2020 German statistics[1]. Each home was assigned a private means for commuting, represented by a car in the virtual world. The car serves as a proxy for commuting by foot or other private means of transportation. A proportion of the population is selected at the beginning of the simulation to commute by bus. Please refer to the ODD protocol for details.

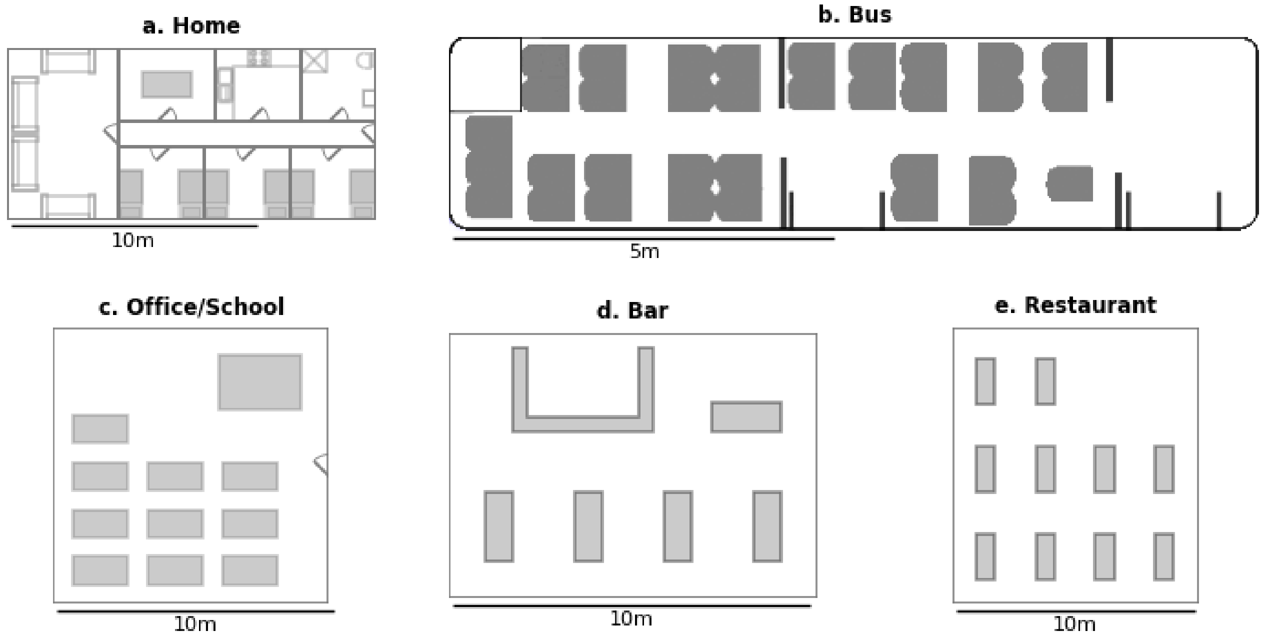


*Fig. S2:* Some of the buildings were given internal structures to add spatial reference for activity execution, including (a) rooms in the home, and tables in the (c) office / classroom, (d) bar, and (e) restaurant, while (b) bus seats were used to spatially arrange agents during rides. Dimensions of all virtual world structures were derived from actual building sizes, and reference material, e.g., public bus manufacturer catalogue.

Each location provides a unique set of activities that can be performed by agents. For simplicity, we restricted activities to locations or to specific furniture, as depicted in Fig. S2. For example, homes were fitted with several subspaces and furniture, allowing agents to engage in different activities, including Resting by sitting on the couch in the living room. Similarly, the activity EatingOut happens when the agent is at the bar or restaurant .

### Validation and calibration

#### Validation of daily activity patterns

To validate activity patterns, we matched daily instances per agent and daily cumulative activity duration per agent with existing datasets. We selected the Extrasensory[2] (ES) dataset as baseline because its labelled activity granularity matched ours. In ES, 60 participants annotated their activities e.g., sleeping, resting, working, for a week. While larger activity recognition repositories exist, e.g., UK biobank[3] with 100,000 participants, and an average 7.6 days of recording, labels are sparse and the granularity limited to individual actions such as typing or drinking from a cup , which are not comparable to our activity abstractions. We simulated 50 independent runs of 1,000 agents, each run covered seven days, without a pathogen model. We created an associative mapping of ES' activities to I2MB activities. For example, lying down (not sleeping), watching tv, and sitting at home labels in ES were mapped to OtherHomeAct activity in I2MB. The complete mapping is available in the source code in simulator/activity_utils/extrasensory.py

In Tab. S1, we present the average daily activity instance count per agent measured in two ways: (1) normalised by including individuals/agents, who actually performed the activity on a given day, and (2) normalised across the total population, which includes those who did not perform the activity. Due to missing activity logging of the participants in ES, frequencies across the total population appear too low (e.g., toilet, less than once per day). Overall, the simulated (I2MB) frequencies across the total population appear reasonable, and a sufficient overlap between ES and I2MB can be observed.

Compared to the normalisation by total population, normalisation by performing individuals/agents showed a better overlap between ES and I2MB. We assume that when ES participants logged an activity, they did so consistently during that day. No variance was observed for CommuteBus, CommuteCar, and EatOut in I2MB, as each commuting agent did so exactly twice per day, and each agent could only use the EatOut activity once daily. The variances observed for the abovementioned activities in the ES study could be associated with changes in commute mode during a day or that locations in cars and buses were logged in more situations than as part of a daily commute. The largest absolute differences were observed for coffee breaks (1.5/d in ES vs. 2.4/d for simulations) and work (3.9/d in ES vs. 3.0/d for simulations).

| Activity | Daily instances per agent [Mean ± SD] | | | |
|---|---|---|---|---|
| | Normalised by performing individuals/agents per day | | Normalised by total population | |
| | ES | I2MB | ES | I2MB |
| **CoffeeBreak** | 1.5 ± 0.8 | 2.4 ± 1.0 | 0.0 ± 0.3 | 2.1 ± 1.3 |
| **CommuteBus** | 1.6 ± 0.8 | 2.0 ± 0.0 | 0.1 ± 0.4 | 0.2 ± 0.6 |
| **CommuteCar** | 2.1 ± 1.3 | 2.0 ± 0.0 | 0.2 ± 0.7 | 1.5 ± 0.8 |
| **Eat** | 2.0 ± 1.2 | 2.0 ± 0.9 | 0.4 ± 1.0 | 1.7 ± 1.1 |
| **EatOut** | 1.3 ± 0.5 | 1.0 ± 0.0 | 0.1 ± 0.3 | 0.0 ± 0.2 |
| **Grooming** | 1.6 ± 0.9 | 1.6 ± 0.5 | 0.1 ± 0.5 | 1.3 ± 0.7 |
| **KitchenWork** | 1.5 ± 0.9 | 2.2 ± 1.1 | 0.1 ± 0.5 | 1.8 ± 1.3 |
| **OtherHomeAct.** | 5.7 ± 6.1 | 5.7 ± 1.9 | 1.2 ± 3.6 | 4.9 ± 2.6 |
| **Shower** | 1.2 ± 0.5 | 1.6 ± 0.5 | 0.1 ± 0.3 | 1.3 ± 0.7 |
| **Sleep** | 1.9 ± 2.2 | 1.2 ± 0.4 | 0.3 ± 1.1 | 0.9 ± 0.6 |
| **Toilet** | 1.7 ± 1.0 | 2.1 ± 1.0 | 0.1 ± 0.6 | 1.7 ± 1.3 |
| **Work** | 3.9 ± 2.7 | 3.0 ± 1.1 | 0.9 ± 2.1 | 2.6 ± 1.4 |

*Tab. S1: Averaged daily activity instances per individual in ES and agent in I2MB. Frequencies were computed (1) normalised by performing individuals/agents on the day, and (2) normalised by the total population size. Normalisation by total population showed that some activities were implausibly annotated in ES, e.g. "Toilet" use less than once per day is unlikely. Normalisation by performing individuals/agents showed reasonable overlap between simulation (I2MB) and annotations (ES). Some differences can be explained by the ES participants' student lifestyle that contrast with the urban worker lifestyle assumed in the simulation.*

We randomly selected 1,000 samples per activity from the daily cumulative activity duration per agent generated from 50 runs of behaviour simulations (total approx. 350,000 samples). Fig. S3 compares daily cumulative activity duration distributions per agent between simulations and ES. The interquartile ranges (IQRs) of simulated activities overlap with their ES counterparts. Distribution differences can be explained by the ES study population and its geographical location, i.e. ES included university students from one city (San Diego, California). In particular, commuting durations were shorter in ES, with a median of 20 min, whereas our simulated commutes had a median duration of 70 min in public transport. Simulated commuting durations were aligned with those of office workers in urban regions sampled from public transportation network route durations[4].

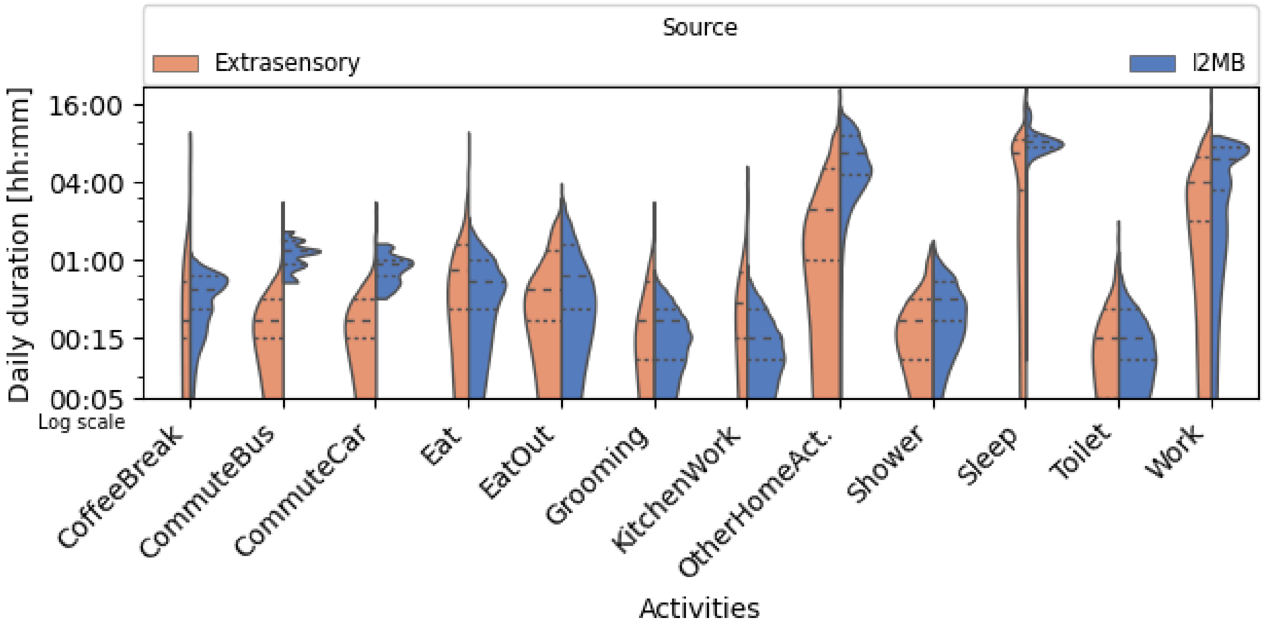


*Fig. S3: Daily cumulative activity duration per agent distributions of I2MB simulations in comparison to Extrasensory (ES) study data. The 25th, 50th (median), and 75th percentiles (*interquartile ranges, IQRs*) are shown for all activities. Distribution differences can be attributed to the specific ES population characteristics. ES mainly comprises on-campus students, where duration of commuting and other activities do not conform to whole-population averages.*

#### Validation of contact patterns

We validated the I2MB simulator's generated contact patterns against Tomori et al.[5], who investigated contact person frequencies in different pandemic studies, referred to as waves. For a definition of a contact person, please see the main document. Repeated contacts with the same person within a given time period count only once. In addition, Tomori et al. considered group contacts as the number of remembered contact persons, where listing them individually was not possible, e.g., clients at a store.

We replicated behaviour patterns of POLYMOD and COVIMOD study conditions[5] by simulating (1) no intervention, (2) closure of bars and restaurants (CBR), and (3) a population fraction staying at home. Each condition was simulated 50 times with 1,000 agents for seven days. For each run, we generated new friends and family networks as well as new house and office assignments. We tracked each agent's daily contact persons and stratified them by locations "Home", "Work", "Transportation", and "Other".

Here we focus on the behaviour of contact persons per location and overall. We did not include age or gender in the simulator model. Unlike Tomori et al., we did not apply any survey or normalisation weights per household size to our simulations since the simulations follow Germany's household size distribution[1]. To compare with Tomori et al., we show both weighted and unweighted data from their result.

NPI restrictions during each COVIMOD wave were extracted from the first, fourth, fifth, and sixth Bavarian Infection Protection Measures Ordinance[6]. For each simulation of a COVIMOD wave, we adjusted the lockdown level by setting the percentage of the population that went to work. The population that had to go to work was split into two groups: fixed and dynamic. The fixed group represents a population share that could not stay at home during daytime. The fixed group was selected at the beginning of each run and had a size of 10% of the total population. The dynamic group was selected every day from the remaining population, with the group size set to 5%, 20%, 40%, and 60% of the total population. Thus, the total percentages of people who went to work were 15%, 30%, 50%, and 70% for COVIMOD waves 1 through 4, respectively. For COVIMOD waves 1 and 2, bars and restaurants were closed. Tab. S2 details parameter combinations for each COVIMOD wave that was replicated by our simulations.

| Study / wave | Study execution date | Restrictions |
|---|---|---|
| POLYMOD | 2005 - 2006 | No restriction |
| COVIMOD 1 | 30/04 to 06/05/2020 | B&R closure, offices 10% fixed, 5% dynamic |
| COVIMOD 2 | 14/05 to 21/05/2020 | B&R closure, offices 10% fixed, 20% dynamic |
| COVIMOD 3 | 28/05 to 04/06/2020 | B&R closure from 22:00, offices 10% fixed, 40% dynamic |
| COVIMOD 4 | 11/06 to 22/06/2020 | B&R closure from 22:00, offices 10% fixed, 60% dynamic |

*Tab. S2: Summary of simulator configurations (I2MB) to replicate the* POLYMOD and COVIMOD studies. *We used* Tomori et al.'s *study execution dates to estimate the closure of bars and restaurants and the proportion of the population allowed to go to work. See text for details on fixed and dynamic population shares.*

We recreated the plots for the weighted and unweighted contacts, with and without group contacts using a negative binomial distribution with parameters derived from the corresponding mean and standard deviation reported in File 3 of Tomori et al.'s supplemental material[5].

Overall, our simulations matched contact person distributions for all study conditions. The largest differences were observed for COVIMOD waves 1 and 2, which can be attributed to differences in average household sizes. For our simulations, the average household size was two residents. In contrast, the sampled population in POLYMOD and COVIMOD studies had an average household size of three. Consequently, in simulations, we observed one contact person on average, while COVIMOD reports two. As restrictions are relaxed in subsequent waves, contact persons increase and non-household contact persons become more relevant.

#### Overall contact patterns

The results show that simulated contact person count increased, as it was observed in Tomori et al.'s results after correcting for group contacts. We find that simulated contact person profiles are within less than Tomori et al.'s reported experiment standard deviation. However, the results also indicate that the virtual world considered here may constrain agent interaction: The overall contact person count is below the reported values by five contact persons on average when considering group contacts and by one contact person when considering only reported contacts as shown in Fig. S4.

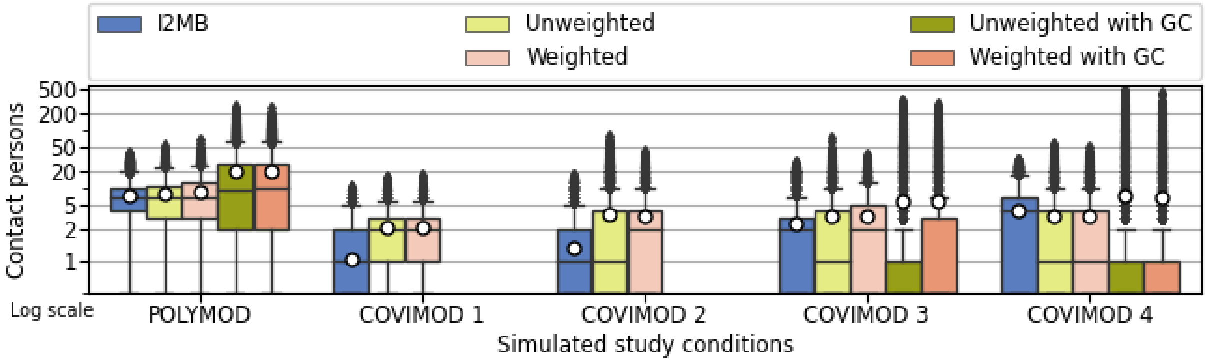


*Fig. S4: Box plots of overall contact person distributions estimated by I2MB simulations and reported by Tomori et al.[5] Boxes illustrate the IQR ([25% to 75%]), line marking the median, and white dots the mean. Whiskers indicate 1.5 times IQR. Average contact person counts generated by I2MB simulations are comparable to contact patterns found by Tomori et al.*

#### Location-based contact patterns

**Location "Home".** Fig. S5 illustrates distributions of contact persons at location home based on I2MB simulations and Tomori et al. We notice that I2MB simulations produced fewer contact persons than reported by Tomori et al., in particular for POLYMOD. Differences can be explained by the lack of friend and family visits in the simulation and the lack of a joint household activity schedule for agents at home, e.g., dinner time. In our simulations, agents determined their home activities individually.

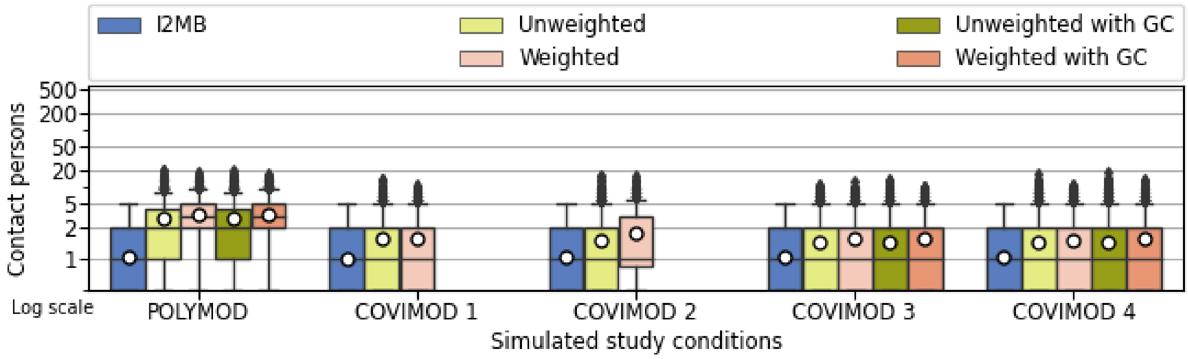


*Fig. S5: Box plot of contact person distributions estimated by I2MB simulations and reported by Tomori et al.[5] at location "home". During the COVIMOD waves, when friends and family visits were suspended, we saw the effect of the joint household schedules and household visits.*

**Location "Work".** Fig. S6 illustrates distributions of contact persons at location work, based on I2MB simulations and Tomori et al. We noticed that I2MB simulation averages reflect the unweighted averages (i.e. without group correction) reported by Tomori et al. Simulations covered 20-person offices only, which may explain why simulations showed smaller IQR than those reported.

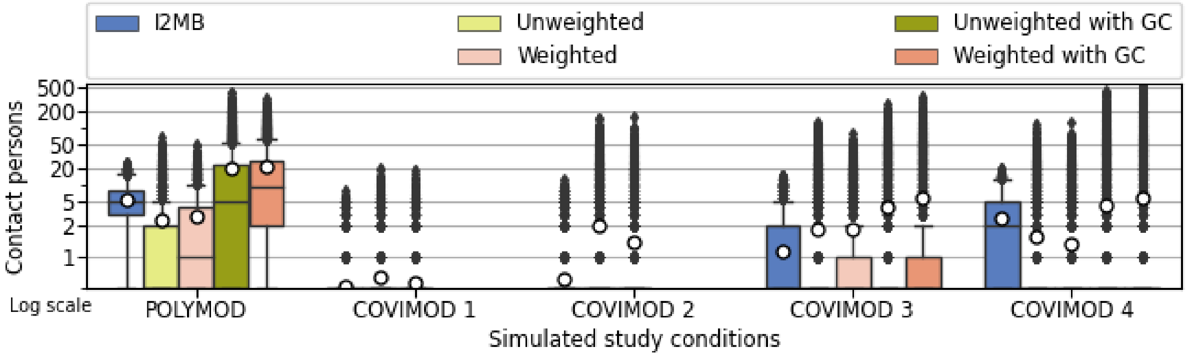


*Fig. S6: Box plot of contact person distributions estimated by I2MB simulations and reported by Tomori et al.[5] at location "work". On average, I2MB simulation results appear similar to unweighted contact person reports. Our simulations covered up to 20-person offices, which may explain the lower IQR compared to those reported by Tomori et al.*

**Location "Transport".** Fig. S7 illustrates distributions of contact persons at location transport, based on I2MB simulations and Tomori et al.The simulations' transportation configuration generates contact patterns that match reported values.

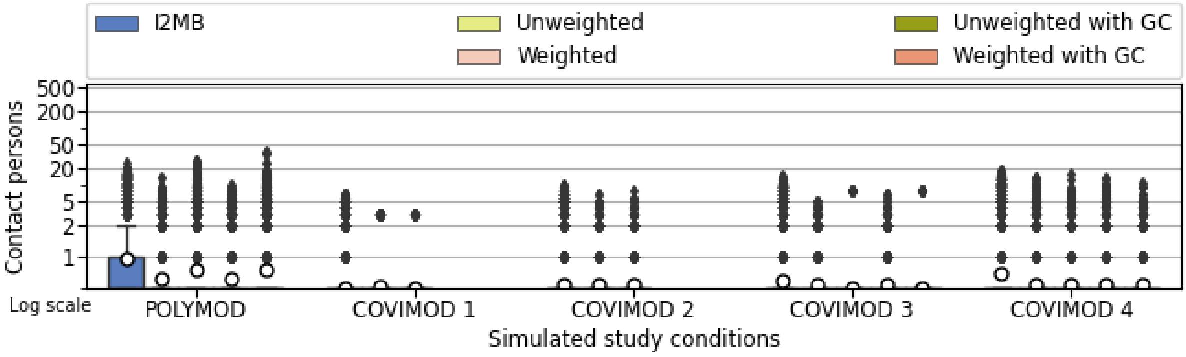


*Fig. S7: I2MB simulated transportation configuration generated contact patterns that match reported values by Tomori et al.*

**Location "Other".** Fig. S8 illustrates distributions of contact persons at location other, based on I2MB simulations and Tomori et al. While restaurant and bar provided agent interactions, the "other" possible contacts were underrepresented. Further work may add other locations and public places, including shops, supermarkets, gyms, and parks.

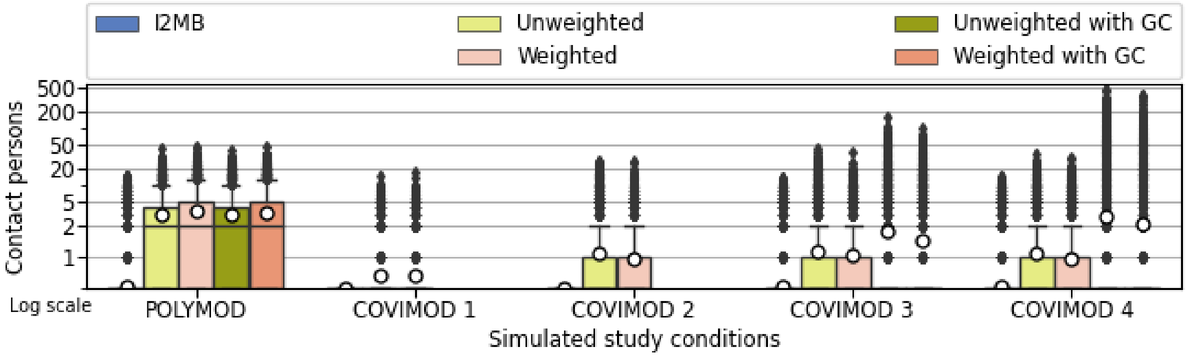

*Fig. S8: Contact person counts reported at other locations. I2MB's simulated contacts in "other" locations are less than one on average for all experiments. We observe that for POLYMOD and COVIMOD 3 and 4, some agents achieve contact persons in the same order of magnitude.*

##### Conclusion on contact pattern validation

The overall contact patterns appeared sufficiently close between I2MB simulations and reports by Tomori et al. Furthermore, location analysis revealed that household, transport, and work contact patterns were comparable, with differences explained by the lack of inter-household visits and the homogeneity of the work location in the simulations. The "Other" location contact patterns differed more widely due to the virtual space limitations. We consider that the virtual world infrastructure always remains a limited abstraction of the real world, even if further spaces would be added. Thus, behaviour in a modelled virtual world is already restricted. If NPIs do not show benefits in the modelled virtual world, they will likely fail in the real world too.

#### Validation and calibration of pathogen characteristics

We introduce a mechanistic pathogen propagation model to describe the exchange of viral load (for detail see methods section in the main paper). To evaluate the pathogen propagation model, we simulated the individual behaviour of 1,000 agents for a full pandemic duration. We show probability density functions (PDFs) of epidemiological variables and compare them with observed COVID-19 statistics. MCT and ICT were included in the simulations to align with the available real world data.

The mechanistic pathogen propagation model has three functional parameters: infectiousness level, exposure function, and viral load removal function. The method section in the main text describes these parameters in detail. Although the model's parameters were based on available evidence, e.g., Kissler et al.[7] We determined the viral load exhaled by an infected individual j as $\frac{1}{6} \cdot il_j(t)$. The exhaled viral load term is included in Eq. 3 of the main manuscript.

Fig. S9 illustrates selected epidemiological variable distributions. Our simulations widely agree with measured distributions for serial[8] and generation[9] intervals. The overall agreement confirms that contact patterns generated by the simulation match natural contact patterns. Furthermore, the simulations agree with reported measurements of incubation[10] and clearance[7] periods, supporting the validity of our novel pathogen propagation model.

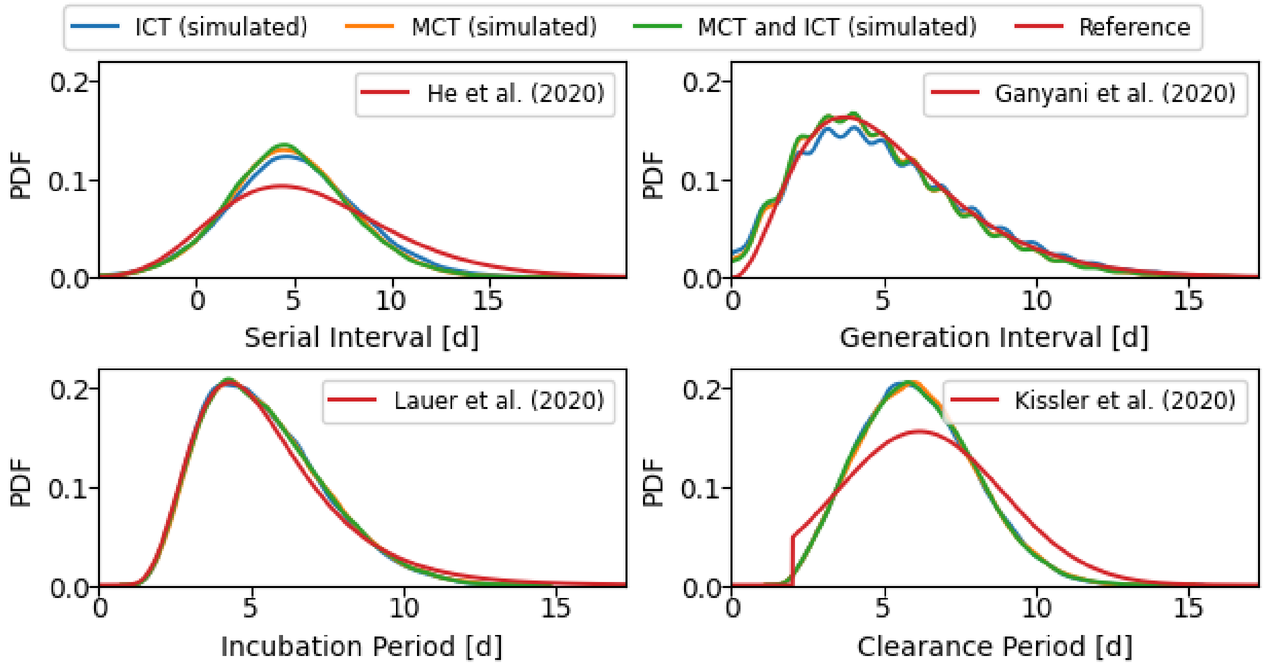


*Fig. S9: Comparison of I2MB simulation and reported epidemiological variables for COVID-19. We assume that ICT and MCT were applied during the reference collection. For the simulations, we varied the presence of ICT and MCT in three simulation configurations, to analyse their impact on the epidemiological variables. Overall, simulation results match the references.* Incubation[10] and clearance[7] *periods were not affected by the analysed contact tracing strategies as they are properties of the interaction between the virus and the individual host. In addition, ICT and MCT did neither have an effect on* serial[8] and generation[9] *intervals.*

#### Network dynamics

As an additional validation step, we calculated the networks created by the interactions between agents and compared them with the unweighted networks used in other agent-based simulators. We defined four contact types in our simulation: with family, friends, acquaintances, and random (See main manuscript [dct_analysis] for details).

For this analysis, we considered four epidemiological concepts with their adapted definitions: (1) contact persons of an agent, regardless whether infectious or not, are the agents within three metres, (2) contacts of an agent are the agent's encounters with contact persons, (3) contact duration is the accumulated encounter duration between two contact persons, and (4) aggregated contact duration of an agent is the accumulated contact duration with all contact persons of that agent. For example, if agent A met with agent B three times for 5 minutes each time and with no one else, the count of agent A's contact persons would be one, the count of contacts would be three, and the contact duration would be 15 minutes. We mapped the epidemiological variables #1 and #4 to network metrics as follows: the count of contact persons (of an agent) maps to the degree (of a node), while the aggregated contact duration maps to the strength (of a node) using the contact duration as weight (on an edge). Eq. S1 defines the degree $d_i$ as the count of edges $E$ that contain node $i$.

$$d_i = |\{\forall e \in E : i \in e\}| \quad \text{(S1)}$$

Eq. S2 illustrates the strength of a node $i$ $s_i$ as the sum of the edge property $p$ for all edges containing $i$.

$$s_i = \sum_{e:\, e \in E} e_p : i \in e \tag{S2}$$

Our analysis is divided into three steps: (1) analyse contact network shapes, (2) correlation between epidemiological variables and network variables, and (3) time series network metrics. Across all, we focus on the network metrics computed using the contact network. We used four scenarios for the analysis: (1) the baseline, where only symptomatic agents isolate, all with compliance of 1.0, (2) MCT and ICT, where people quarantine based on information from friends and families or manual contact tracing following the realistic scenario conditions (see main manuscript for details), (3) realistic behaviour scenario including DCT, where DCT adoption, adherence, and compliance were set to 0.5, 0.7, and 0.7 respectively, and (4) DCT-optimal behaviour scenario, where DCT, MCT and ICT are used to inform contact persons of positive cases, and adoption, adherence, and compliance were all set to 1.0.

#### Contact network shapes

The behavioural approach employed in this study yields contact networks with similar shapes to those generated by agent-based simulators[11]. However, the resulting contact networks exhibit the emergent property of being independently weighted. In contrast, the top-down approach used in other agent-based simulators necessitates data that may not be readily accessible for configuring the weights of each potential network during setup. Fig. S10 illustrates the heterogeneity of networks per contact type, along with the corresponding weights indicated as strengths based on contact duration.

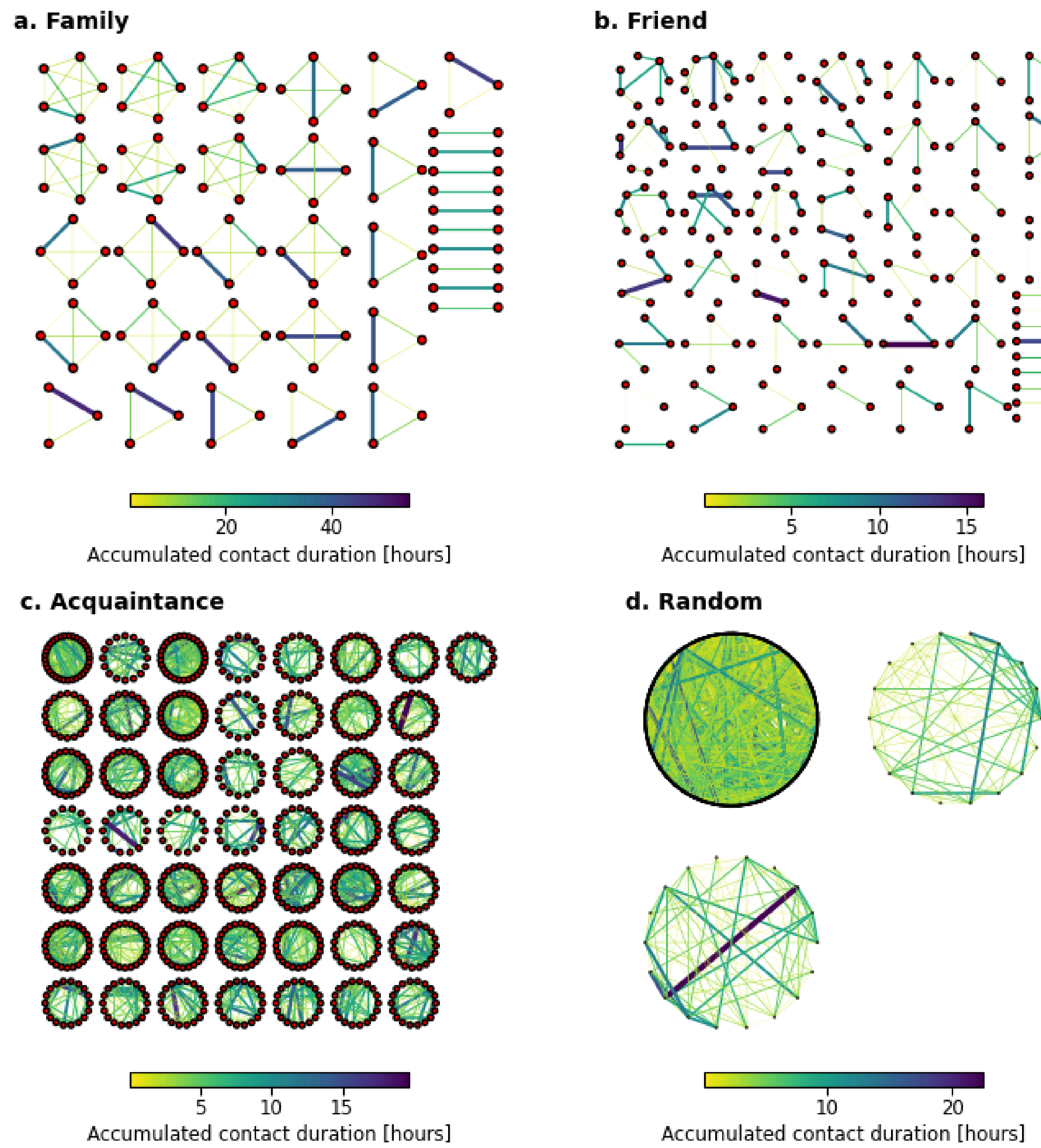


*Fig. S10: Examples of emerging weighted networks by contact type. These examples come from a simulated week without pathogen presence. For each edge we used the accumulated contact duration property to control the width and shading. The generated behaviour driven encounters create heterogeneous weighted networks, illustrating agent interaction preference. The emergent networks have similar shapes to networks used for implementing traditional ABMs, which supports the realism of the behaviour driven simulation paradigm.*

The differences between min-max normalised network degree and strength distributions for different simulation configurations are illustrated in Fig. S11. The degree focuses on contact opportunities, as seen in the bimodal distribution for the random network. For instance, full-time workers are more likely to experience random contacts during their work hours and coffee breaks, public transport commuters during their public transport rides, and diners and patrons when visiting bars and restaurants. In contrast, network strength tracks the cumulative contact duration, which is an unimodal distribution with no discernible groups.

.

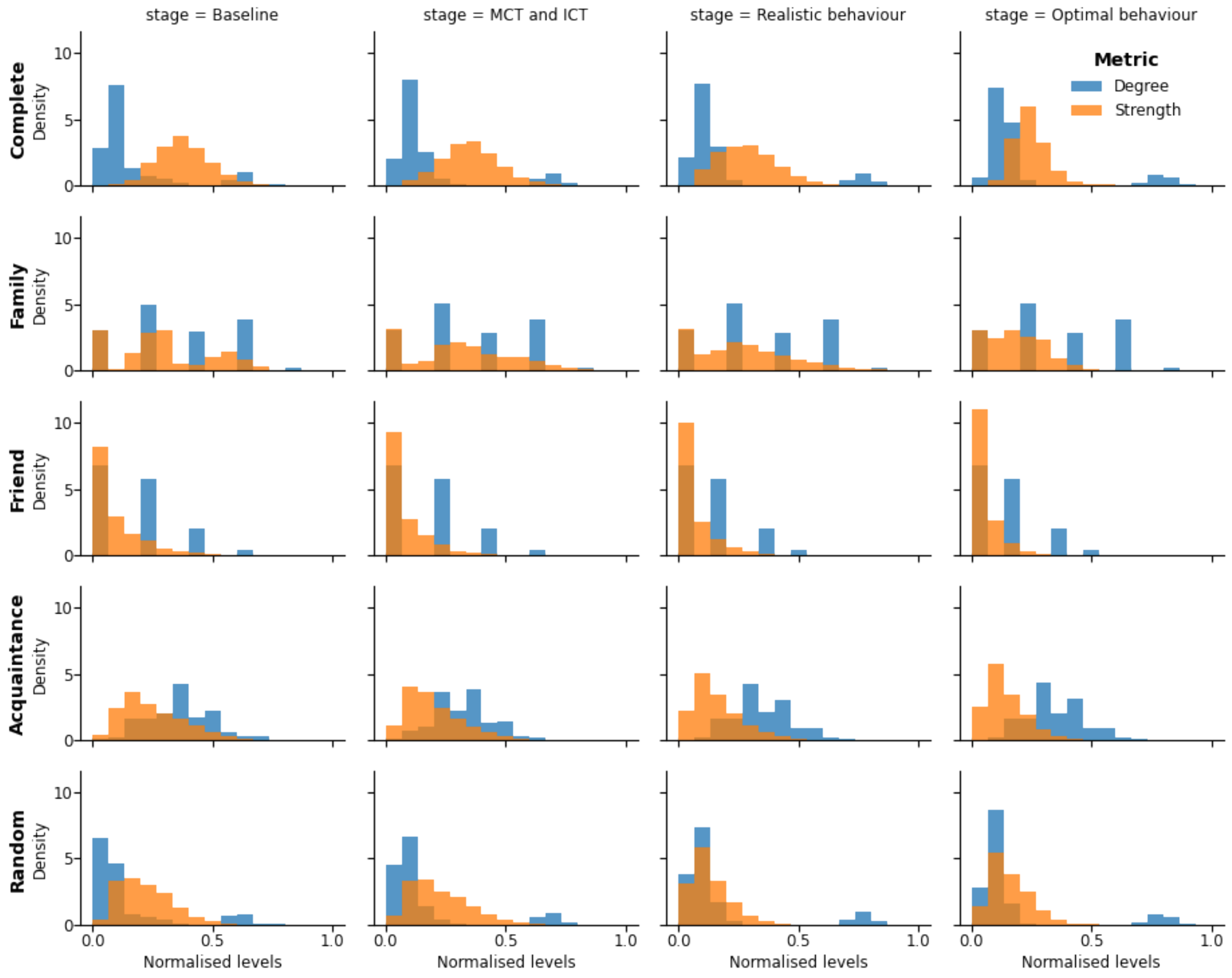


*Fig. S11: Differences between min-max normalised degree and strength distributions, by contact tracing modality and behaviour scenario and by contact type. The degree focuses on contact opportunities, as shown by the multimodality of random networks, where full-time workers and public transport commuters drive the second mode. Strength focuses on the contact duration, follows an unimodal distribution and provides no discernible grouping.*

The lack of connection in the friends' network may be attributed to limited availability of leisure spaces and no inter-household home visits. In future work, we will consider more public spaces, including locations for leisure activities, e.g., parks and malls. In addition, we will enable inter-household visits.

The difference between distributions might lead to different risk assessments and planned interventions. Based on the simulation results, if the degree is used, i.e., unweighted networks, full-time workers, public transport commuters, and gastronomy services, would constitute a target group for contact reduction interventions. In contrast, network strength yields more holistic intervention strategies that would not target any specific social group.

#### Correlation between network metrics and pandemic characteristics

We investigate the relationship between network metrics — specifically degree and duration-weighted strength — and pandemic characteristics such as infection time and 7-day incidence. Our analysis focuses on the infection history, comparing the degree and strength of the infector (source) and the infectee (target) in relation to pandemic progression. As shown in **Fig. S12**, combining different types of contact tracing pushes contact persons from random to family-based networks. In addition, strength shows a stronger correlation to infection time, especially with digital contact tracing (DCT), highlighting the importance of measuring risk based on contact duration rather than contact person counts.

Further analysis (**Fig. S13**) shows that strength correlates more strongly with infection time when measured only before infection, but this insight does not yield actionable prevention strategies as agents infected later in the pandemic have had more time to accumulate strength. When contact metrics are limited to the week preceding infection, the correlation with infection time diminishes (**Fig. S14**), suggesting that socially active agents drive the pandemic, even with small social circles. Moreover, as DCT improves, source agents with larger social networks and longer contact durations drive later-stage infections. The relationship between 7-day incidence and network metrics (**Fig. S15**) reveals a positive correlation between the source's degree and strength and the 7-day incidence, while targets show a negative correlation, especially under optimal DCT conditions. This indicates that low-incidence periods pose higher infection risks for susceptible agents due to increased interaction duration and the availability of infectious agents. The findings suggest that future DCT implementations could enhance pandemic control by encouraging individuals to monitor and moderate strength, i.e., cumulative contact duration, particularly during periods of low incidence.

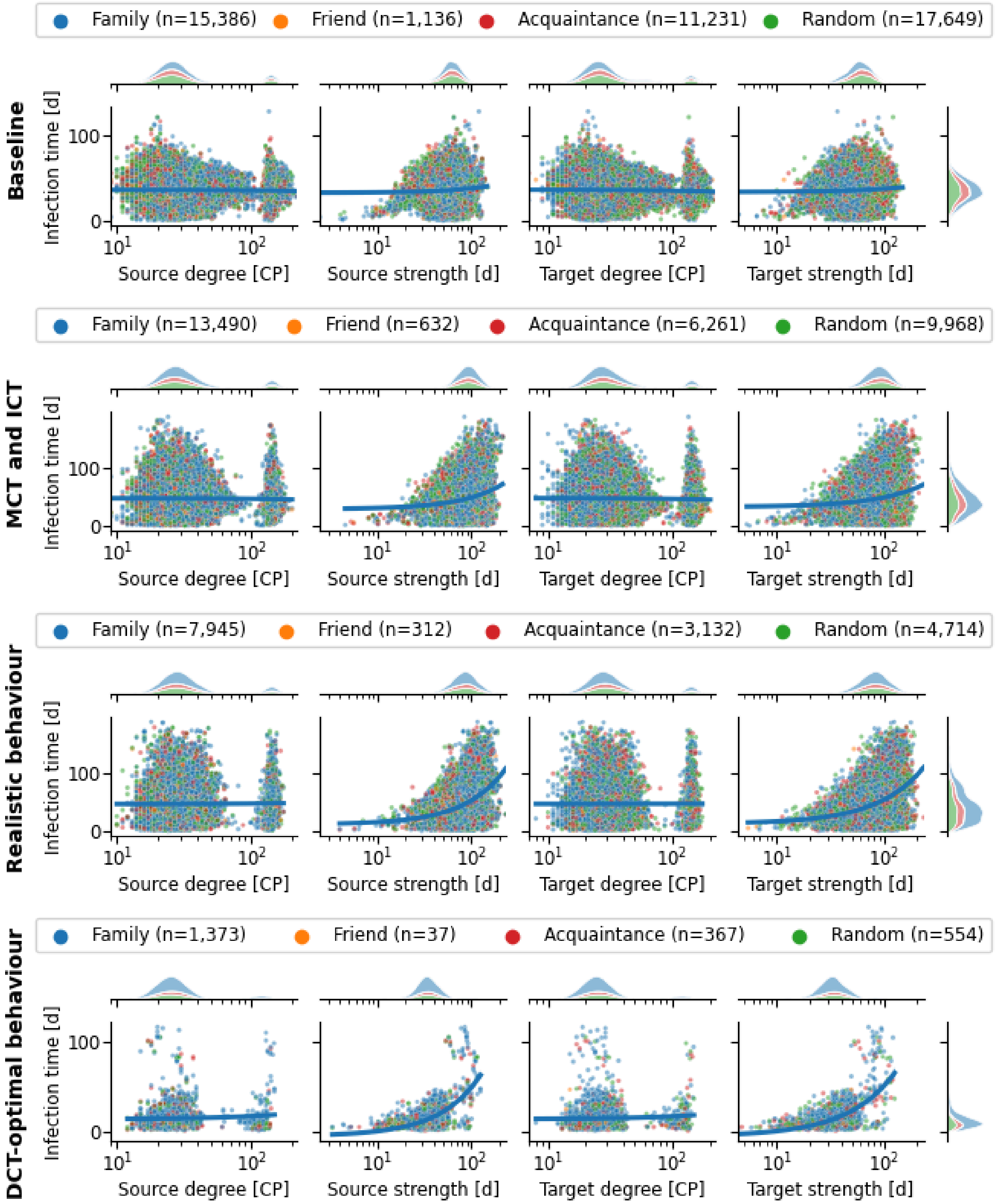


*Fig. S12: Illustration of overall degree and strength, which are measured during the entire simulation, vs. infection time. Overall degree (contact persons [CP]) of the source and target are not correlated to the infection time. Overall strength (aggregated contact duration in days [d]) shows a positive correlation in each of the test scenarios.*

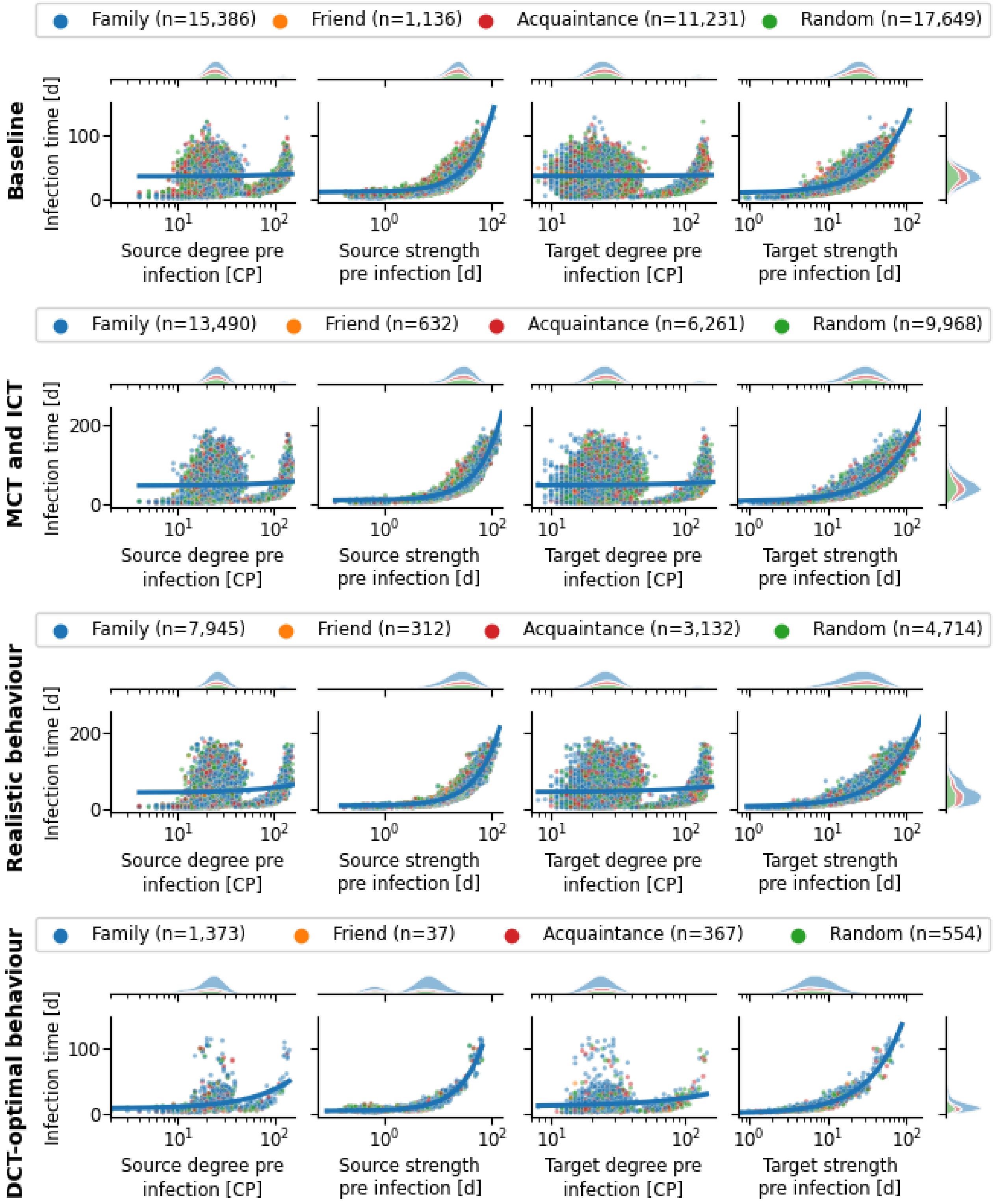


*Fig. S13: Degree (contact persons [CP]) and strength (aggregated contact duration in days [d]) before the target's infection time. The improved correlation between strength and infection time yields no actionable insight, as the target has had more time to accumulate connections.*

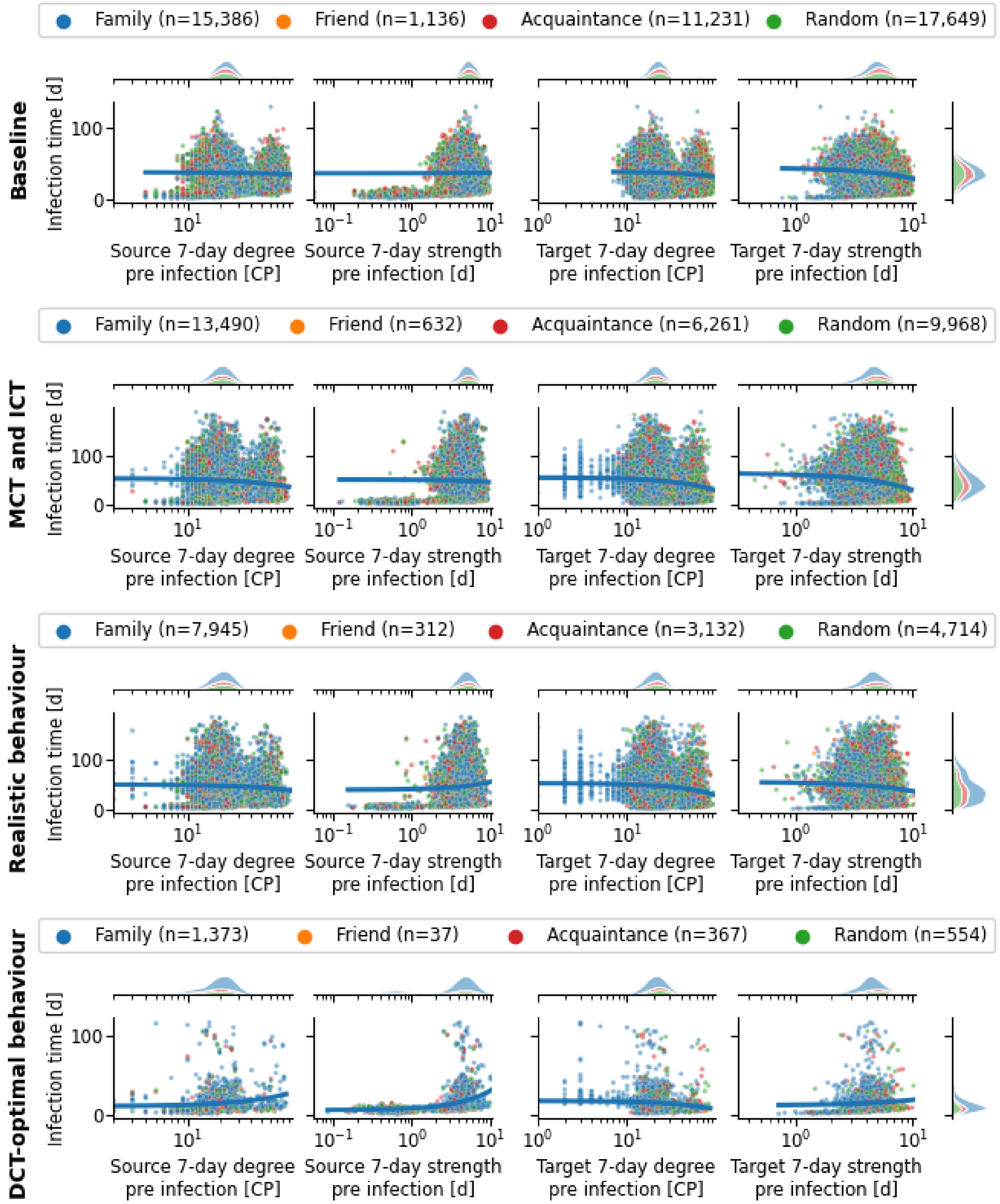


*Fig. S14: Degree (contact persons [CP]) and strength (aggregated contact duration in days [d]) measured during the 7-day period prior to the target's infection. Correlations between strength and infection time collapse when considering the contacts during the 7-day prior to the target's infection. The negative correlation with the target degree indicates that the pandemic will progress even if the target has small social groups.*

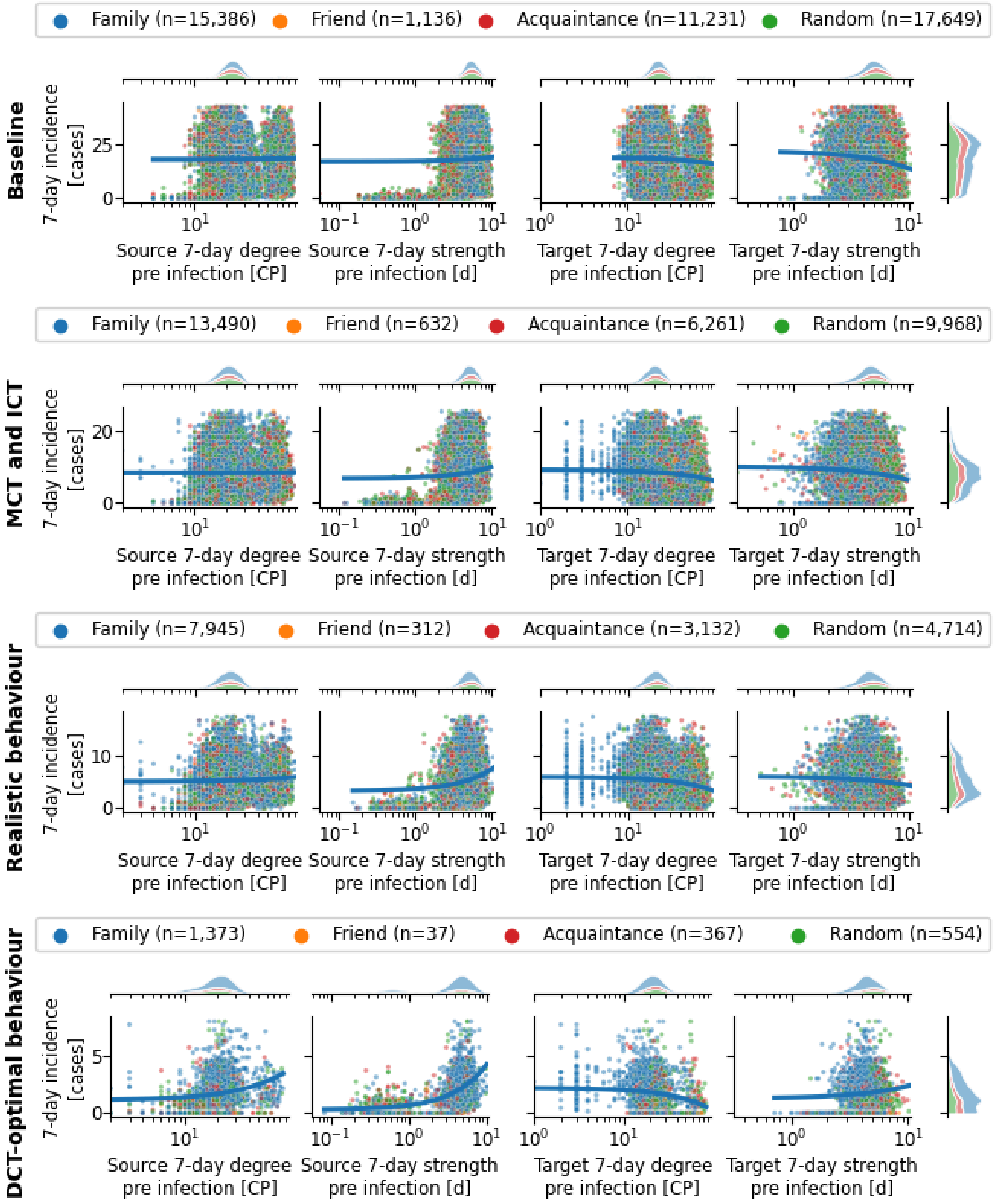


*Fig. S15: Degree (contact persons [CP]) and strength (aggregated contact duration in days [d]) measured during the 7-day period prior to the target's infection. The source degree and strength follow the intuition that a socially active source will cause more infections. The negative correlation of the target's degree and strength indicates that social agents are more at risk when the incidence is low at infection time.*

#### Analysis of daily contacts, degree, and strength

In this section, we examine the daily trends of contact metrics—contacts, degree, and strength—across four different scenarios. **Fig. S16** highlights the baseline scenario without contact tracing, where contacts, degree, and strength remain largely stable throughout the pandemic. A slight increase in all metrics occurs the day before infection, followed by a small reduction in acquaintances and random contacts as individuals self-isolate. However, no shift toward increased family contact persons is observed during isolation in this scenario. Introducing manual (MCT) and individual contact tracing (ICT), as shown in **Fig. S17**, leads to a decline in daily contacts, degree, and strength as the pandemic progresses, returning to pre-pandemic levels by the end of the simulation. Relative time plots reveal that family interactions intensify during quarantine, while contact persons with acquaintances and random individuals decline, aligning with earlier findings that quarantine heightens the risk of household transmission.

In scenarios with digital contact tracing (DCT), MCT, and ICT, **Fig. S18** shows a moderate reduction in random contact persons as isolation and quarantine measures take effect, which is not seen in the baseline scenario where all metrics remained constant. The relative time plots show that contact tracing effectively reduces the daily degree after infection, although family degree remains steady, and family strength increases due to home isolation. **Fig. S19** demonstrates the impact of quarantine over time, with external contact persons decreasing and family interactions increasing during the first 14 days of the pandemic. As individuals leave isolation after 20 days, metrics begin to normalize, although a secondary reduction in contacts is observed after 90 days. Even under optimal DCT conditions, not all infection chains are broken in the first 20 days. Notably, across all scenarios, there is a slight increase in contacts, degree, and strength on the day before infection, suggesting future work could explore contact proximity as a factor to further refine contact tracing strategies.

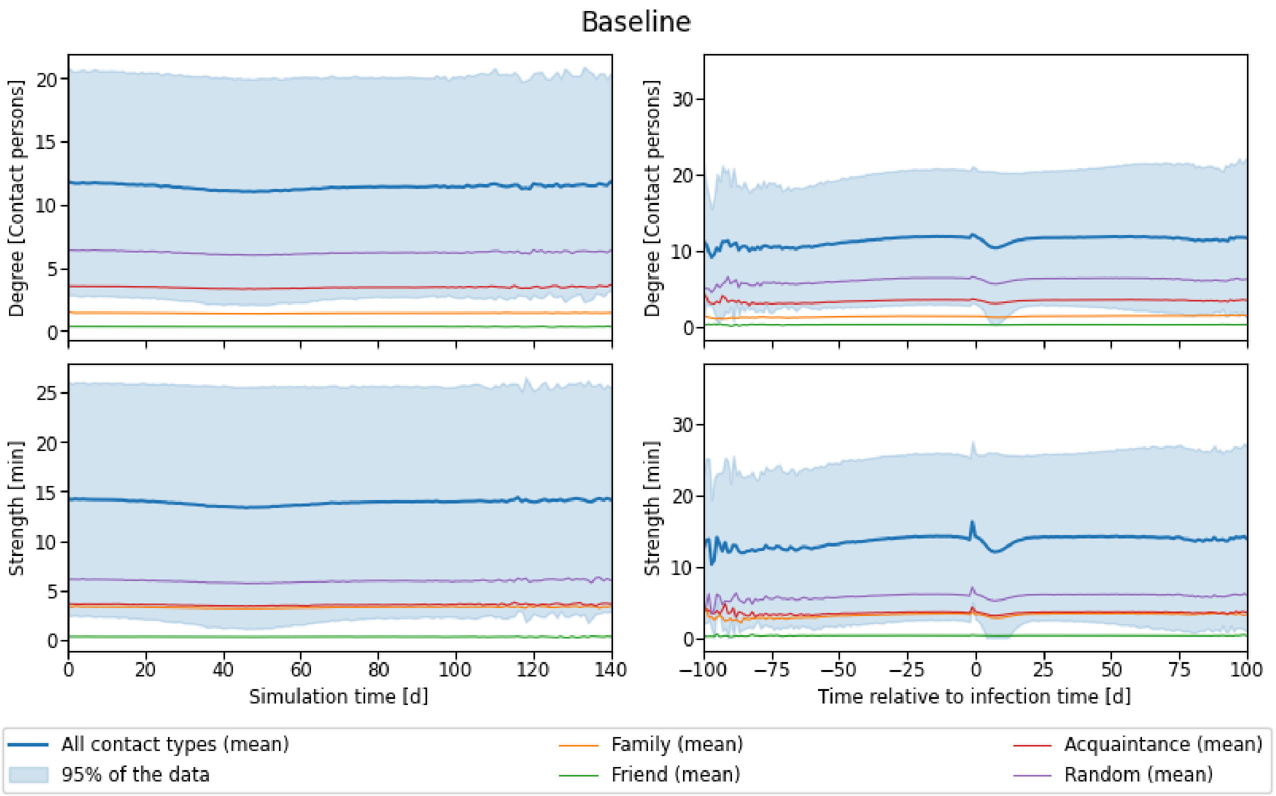


*Fig. S16: Time series data for daily contacts, degree (contact persons), and strength (aggregated contact duration) for the baseline scenario and discarding hospital contacts. Strength is also constant throughout the simulations after removing hospital contacts. A small peak in contacts, degree and strength just before the infection remains.*

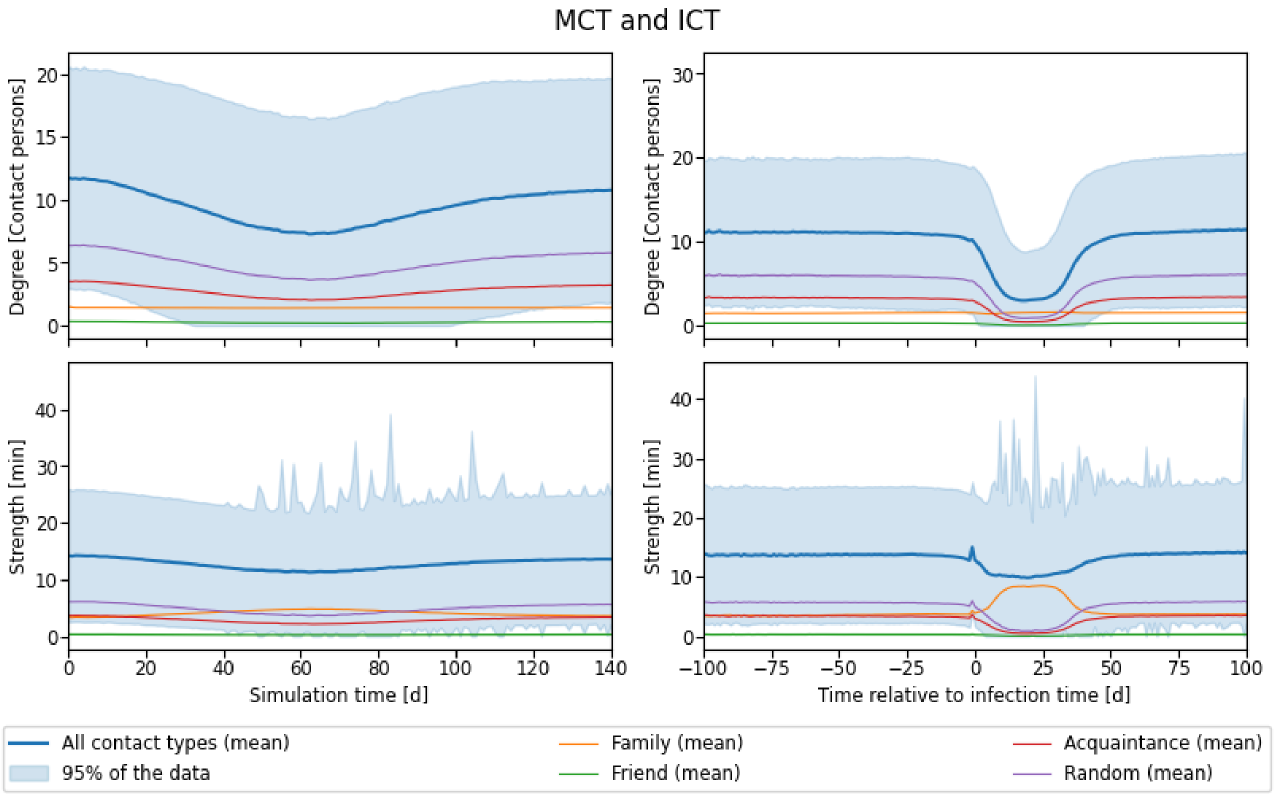


*Fig. S17: Time series data for daily contacts, degree (contact persons), and strength (aggregated*

*contact duration) for the MCT and ICT scenario and discarding hospital contacts. While contacts and degree show a considerable reduction in overall contacts, only strength shows the increased interaction with household members.*

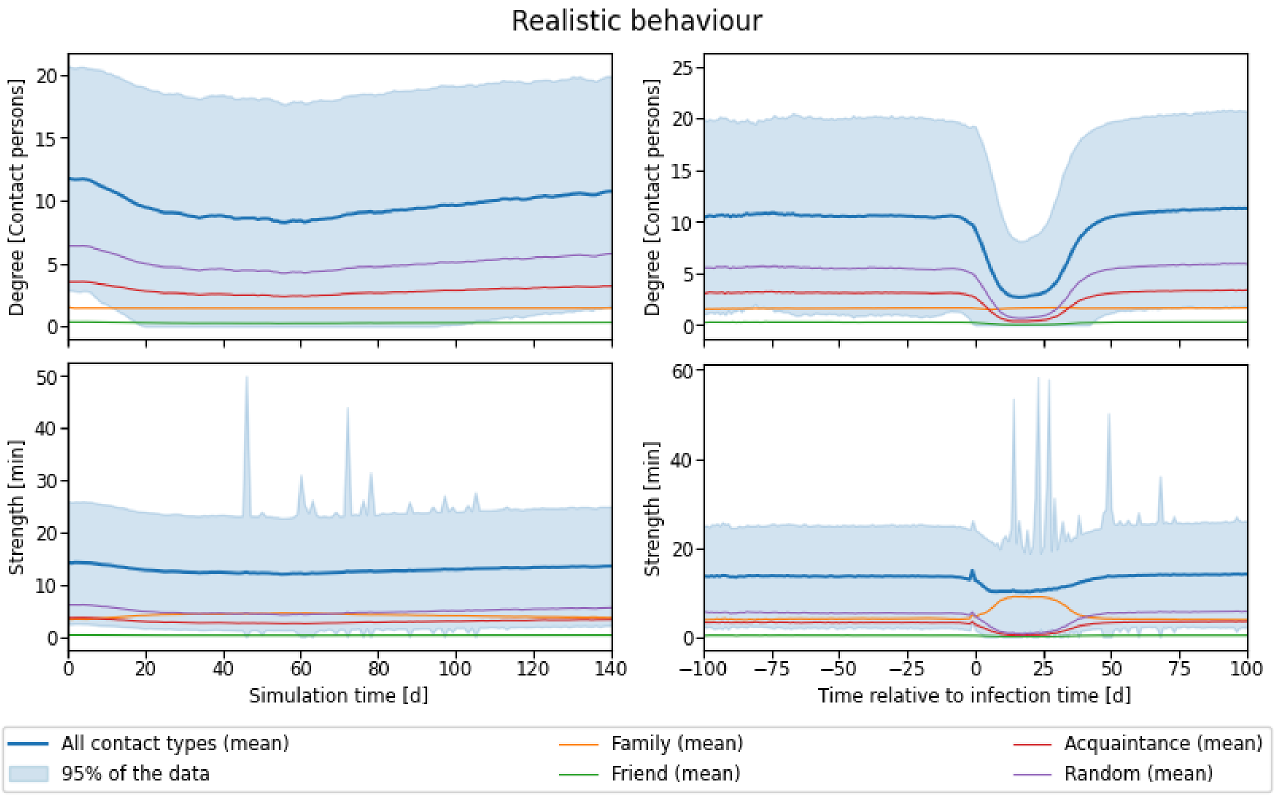


*Fig. S18: Time series data for daily contacts, degree (contact persons), and strength (aggregated contact duration) for the realistic behaviour scenario and discarding hospital contacts. In the realistic behaviour scenario, we observed a faster degree reduction after infection when compared to MCT and ICT, which highlights the impact of using DCT, as the degree restoration happens at the same rate. Contacts and strength illustrate how family encounters substitute other types of contact during confinement.*

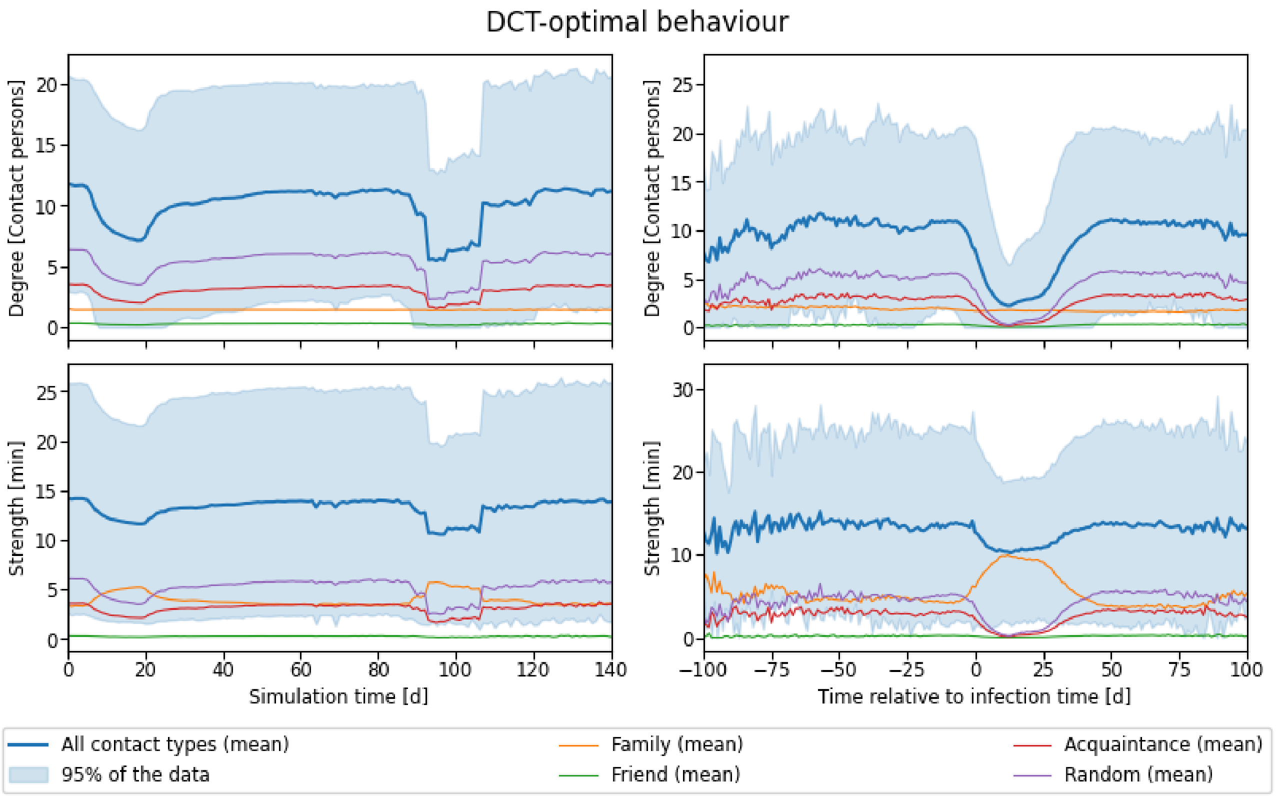


*Fig. S19: Time series data for daily contacts, degree (contact persons), and strength (aggregated contact duration) for the DCT-optimal behaviour scenario and discarding hospital contacts. The simulation time plots show how degree and contacts reduce as positive tests arrive. The time series shows a second wave that occurs, as not all infection chains can be broken.*

#### False discovery rate and false negative rate

| DCT | RNT | QCH | CBR | False discovery rate | | False negative rate | | Fowlkes mallows index | |
|---|---|---|---|---|---|---|---|---|---|
| | | | | Median | IQR | Median | IQR | Median | IQR |
| NO DCT | NO RNT | NO QCH | NO CBR | 88.4% | [88.1%, 88.7%] | 29.1% | [27.8%, 30.1%] | 28.7% | [28.3%, 29.1%] |
| DCT | NO RNT | NO QCH | NO CBR | 97.3% | [96.3%, 97.9%] | 24.8% | [21.5%, 28.2%] | 14.2% | [12.4%, 16.3%] |
| | | | CBR | 97.5% | [96.8%, 98.3%] | 24.5% | [19.5%, 28.9%] | 13.6% | [11.3%, 14.9%] |
| | | QCH | NO CBR | 97.6% | [97.0%, 98.3%] | 24.2% | [20.5%, 26.1%] | 13.5% | [11.5%, 15.0%] |
| | | | CBR | 97.6% | [96.6%, 98.4%] | 24.2% | [19.5%, 27.8%] | 13.9% | [11.3%, 15.8%] |
| | RNT | NO QCH | NO CBR | 93.4% | [91.3%, 94.9%] | 35.9% | [33.2%, 38.9%] | 20.2% | [18.6%, 23.6%] |
| | | | CBR | 92.4% | [90.6%, 94.1%] | 34.4% | [32.2%, 37.9%] | 21.5% | [20.2%, 24.5%] |
| | | QCH | NO CBR | 92.2% | [90.4%, 95.0%] | 35.7% | [32.4%, 38.7%] | 21.8% | [19.1%, 24.8%] |
| | | | CBR | 92.8% | [90.7%, 94.5%] | 35.9% | [31.5%, 38.2%] | 22.2% | [19.6%, 24.0%] |

*Tab. S3: False discovery rate and false negative rate for the NPI combinations in the DCT-optimal behaviour scenarios. The selected scenarios provide a performance boundary for the pandemic characteristics under study, focusing on using DCT as the primary contributor to performance improvement.*

| DCT | RNT | QCH | CBR | False discovery rate | | False negative rate | | Fowlkes mallows index | |
|---|---|---|---|---|---|---|---|---|---|
| | | | | Median | IQR | Median | IQR | Median | IQR |
| NO DCT | NO RNT | NO QCH | NO CBR | 88.0% | [87.7%, 88.6%] | 29.5% | [28.3%, 30.7%] | 28.9% | [28.3%, 29.5%] |
| | | | CBR | 88.3% | [87.9%, 88.7%] | 29.4% | [28.2%, 30.7%] | 28.7% | [28.3%, 29.3%] |
| | | QCH | NO CBR | 87.9% | [87.5%, 88.5%] | 29.2% | [27.2%, 30.1%] | 29.2% | [28.4%, 29.6%] |
| | | | CBR | 88.2% | [87.7%, 88.8%] | 29.6% | [28.1%, 30.7%] | 28.8% | [28.1%, 29.3%] |
| | RNT | NO QCH | NO CBR | 62.7% | [61.7%, 63.9%] | 32.1% | [31.2%, 32.8%] | 50.5% | [49.6%, 51.1%] |
| | | | CBR | 62.9% | [61.6%, 63.8%] | 31.5% | [30.5%, 32.8%] | 50.5% | [49.4%, 51.3%] |
| | | QCH | NO CBR | 63.1% | [61.7%, 64.3%] | 32.1% | [31.1%, 33.3%] | 50.0% | [49.0%, 50.8%] |
| | | | CBR | 62.9% | [61.6%, 64.9%] | 32.0% | [30.9%, 33.0%] | 50.3% | [48.6%, 51.0%] |
| DCT | NO RNT | NO QCH | NO CBR | 92.6% | [92.2%, 93.7%] | 27.5% | [25.6%, 28.5%] | 23.3% | [21.5%, 23.7%] |
| | | | CBR | 93.0% | [92.2%, 94.1%] | 26.5% | [24.3%, 27.6%] | 22.7% | [20.8%, 24.0%] |
| | | QCH | NO CBR | 93.1% | [92.4%, 94.1%] | 27.1% | [25.5%, 28.2%] | 22.5% | [20.8%, 23.4%] |
| | | | CBR | 92.8% | [92.1%, 93.4%] | 26.8% | [25.1%, 28.8%] | 23.1% | [21.7%, 24.1%] |
| | RNT | NO QCH | NO CBR | 76.4% | [75.1%, 78.9%] | 32.0% | [30.7%, 33.2%] | 39.9% | [37.9%, 41.2%] |
| | | | CBR | 76.6% | [75.0%, 78.1%] | 32.3% | [30.8%, 34.1%] | 39.9% | [37.8%, 41.1%] |
| | | QCH | NO CBR | 76.5% | [74.6%, 78.2%] | 31.8% | [30.3%, 33.3%] | 40.3% | [38.4%, 41.6%] |
| | | | CBR | 76.1% | [74.5%, 78.6%] | 32.2% | [30.1%, 33.3%] | 40.2% | [37.9%, 41.8%] |

*Tab. S4: False discovery rate and false negative rate for the different NPI combinations in the realistic behaviour scenarios.*

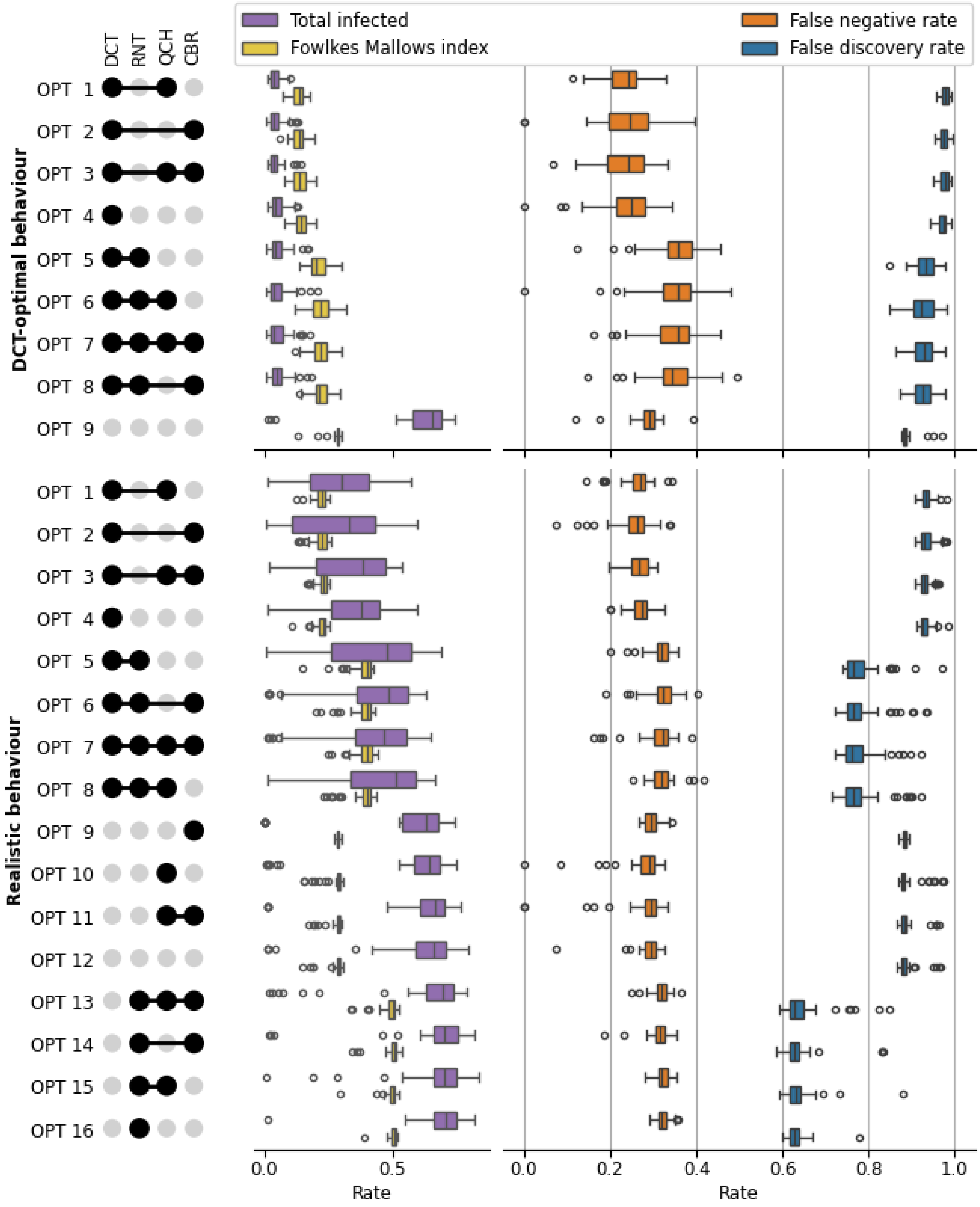


*Fig. S20: False discovery rate and false negative rate distributions for the NPI combinations in the DCT-optimal and realistic behaviour scenarios ordered by the mean total infections with lowest total infections at the top. DCT's success is derived from a low false negative rate (presumably by a slightly faster isolation) at the price of a high false discovery rate, which translates to targeted quarantines that effectively diminishes the risk of infection for highly exposed agents. Note that the best performance based on the total infected epidemiological metric occurs at the worst values for conventional performance metric of classification algorithms, i.e., low Fowlkes Mallows index, due to a high false discovery rate.*

#### DCT-optimal behaviour

| DCT | RNT | QCH | CBR | Generation interval Median | Generation interval IQR | Serial interval Median | Serial interval IQR | 7-day incidence Median | 7-day incidence IQR | 7-day hosp. incidence Median | 7-day hosp. incidence IQR | Total infected Median | Total infected IQR | Wave duration Median | Wave duration IQR | Days in quarantine Median | Days in quarantine IQR |
|---|---|---|---|---|---|---|---|---|---|---|---|---|---|---|---|---|---|
| NO DCT | NO RNT | NO QCH | NO CBR | 4.9 | [3.1, 6.9] | 5.1 | [3.1, 7.2] | 13.2 | [12.0, 16.2] | 3.2 | [2.7, 3.7] | 651.5 | [578.0, 689.5] | 121.2 | [104.4, 137.6] | 24.7 | [21.8, 28.0] |
| DCT | NO RNT | NO QCH | NO CBR | 4.5 | [3.0, 6.3] | 4.7 | [2.8, 6.7] | 2.9 | [2.3, 3.9] | 0.6 | [0.4, 0.9] | 43.5 | [30.0, 65.8] | 36.1 | [31.3, 41.8] | 14.0 | [14.0, 14.0] |
| | | | CBR | 4.6 | [2.9, 6.5] | 4.8 | [2.7, 6.7] | 2.7 | [1.8, 3.7] | 0.6 | [0.4, 0.9] | 35.0 | [25.2, 55.8] | 31.9 | [28.7, 39.7] | 14.0 | [14.0, 14.0] |
| | | QCH | NO CBR | 4.6 | [2.9, 6.5] | 4.7 | [2.5, 6.8] | 2.9 | [2.0, 3.6] | 0.7 | [0.3, 1.0] | 38.0 | [24.0, 54.8] | 33.2 | [30.4, 38.9] | 14.0 | [14.0, 14.0] |
| | | | CBR | 4.2 | [2.8, 6.0] | 4.7 | [2.7, 6.7] | 2.7 | [2.0, 4.0] | 0.7 | [0.4, 1.0] | 39.0 | [25.2, 50.0] | 33.5 | [29.7, 37.4] | 14.0 | [14.0, 14.0] |
| | RNT | NO QCH | NO CBR | 4.4 | [3.0, 6.4] | 4.8 | [2.8, 6.8] | 2.9 | [2.0, 3.8] | 0.7 | [0.5, 0.9] | 42.0 | [28.5, 65.0] | 34.0 | [29.3, 41.5] | 4.1 | [4.0, 5.0] |
| | | | CBR | 4.4 | [2.9, 6.3] | 4.9 | [2.8, 6.9] | 2.9 | [2.2, 4.8] | 0.8 | [0.4, 1.1] | 49.0 | [30.5, 67.5] | 36.1 | [30.9, 48.1] | 4.1 | [4.0, 5.0] |
| | | QCH | NO CBR | 4.3 | [2.9, 6.1] | 4.7 | [2.7, 6.7] | 2.8 | [1.9, 4.1] | 0.7 | [0.6, 1.0] | 39.5 | [24.5, 68.8] | 34.1 | [28.2, 44.9] | 4.1 | [4.0, 5.1] |
| | | | CBR | 4.6 | [2.9, 6.4] | 4.8 | [2.8, 6.8] | 2.6 | [2.0, 3.4] | 0.6 | [0.3, 1.0] | 39.5 | [25.2, 69.5] | 35.4 | [27.8, 55.8] | 4.1 | [4.0, 5.0] |

*Tab. S5: Median and IQR of pandemic characteristics for DCT-optimal behaviour. This table complements Fig. 3a from the main document. In DCT's optimal behaviour, we observe that DCT is the main contributor to the reduction of the pandemic characteristics. The only clear differentiator is the use of DCT and RNT and its effect on days in quarantine. For other combinations, the IQRs overlap indicates that additional pandemic characteristics might be necessary when selecting NPI combinations. The highlighted rows correspond to the selected baselines from FIg 3.*

#### Realistic behaviour

| DCT | RNT | QCH | CBR | Generation interval Median | Generation interval IQR | Serial interval Median | Serial interval IQR | 7-day incidence Median | 7-day incidence IQR | 7-day hosp. incidence Median | 7-day hosp. incidence IQR | Total infected Median | Total infected IQR | Wave duration Median | Wave duration IQR | Days in quarantine Median | Days in quarantine IQR |
|---|---|---|---|---|---|---|---|---|---|---|---|---|---|---|---|---|---|
| NO DCT | NO RNT | NO QCH | NO CBR | 4.9 | [3.1, 6.9] | 5.1 | [3.1, 7.2] | 15.7 | [12.4, 17.7] | 3.6 | [2.9, 4.1] | 660.0 | [592.0, 708.0] | 107.3 | [94.0, 130.8] | 24.6 | [22.0, 27.5] |
| | | | CBR | 4.9 | [3.1, 7.0] | 5.2 | [3.1, 7.3] | 11.9 | [10.0, 14.4] | 2.9 | [2.3, 3.6] | 633.0 | [538.5, 679.8] | 122.6 | [107.9, 144.1] | 24.7 | [22.1, 28.0] |
| | | QCH | NO CBR | 4.9 | [3.1, 7.0] | 5.2 | [3.1, 7.3] | 14.4 | [11.4, 18.0] | 3.4 | [2.8, 3.9] | 641.0 | [582.0, 684.5] | 99.9 | [89.7, 120.9] | 24.6 | [22.0, 27.3] |
| | | | CBR | 4.9 | [3.1, 7.0] | 5.2 | [3.1, 7.3] | 13.6 | [10.7, 17.7] | 3.1 | [2.6, 4.0] | 665.5 | [605.5, 699.8] | 119.9 | [98.6, 138.2] | 24.7 | [22.0, 28.0] |
| | RNT | NO QCH | NO CBR | 4.9 | [3.1, 6.9] | 5.2 | [3.1, 7.3] | 16.2 | [14.0, 19.5] | 3.8 | [3.1, 4.5] | 707.0 | [657.5, 749.0] | 109.3 | [100.6, 123.6] | 5.0 | [2.8, 8.7] |
| | | | CBR | 4.9 | [3.1, 6.9] | 5.2 | [3.1, 7.2] | 16.5 | [14.6, 18.5] | 3.7 | [3.1, 4.4] | 700.5 | [663.0, 752.2] | 112.9 | [97.7, 126.3] | 5.0 | [2.8, 8.7] |
| | | QCH | NO CBR | 4.9 | [3.1, 6.9] | 5.1 | [3.1, 7.2] | 16.3 | [13.8, 19.7] | 3.6 | [3.0, 4.4] | 699.5 | [661.0, 747.8] | 118.0 | [103.4, 132.6] | 4.9 | [2.8, 8.8] |
| | | | CBR | 4.9 | [3.1, 6.9] | 5.2 | [3.1, 7.3] | 14.8 | [11.2, 18.8] | 3.3 | [2.8, 4.3] | 695.5 | [629.5, 739.5] | 118.5 | [101.5, 133.0] | 4.9 | [2.9, 8.6] |
| DCT | NO RNT | NO QCH | NO CBR | 4.8 | [3.1, 6.8] | 5.0 | [2.9, 7.1] | 8.6 | [5.9, 10.1] | 2.0 | [1.6, 2.4] | 379.5 | [260.8, 451.0] | 109.3 | [79.9, 130.9] | 24.9 | [14.0, 35.5] |
| | | | CBR | 4.8 | [3.0, 6.8] | 5.0 | [3.0, 7.1] | 8.3 | [5.2, 10.6] | 1.8 | [1.2, 2.4] | 328.5 | [109.8, 432.5] | 93.2 | [55.7, 125.5] | 24.3 | [14.0, 28.0] |
| | | QCH | NO CBR | 4.8 | [3.0, 6.8] | 5.0 | [3.0, 7.2] | 7.2 | [4.8, 9.8] | 1.7 | [1.3, 2.3] | 300.0 | [176.8, 410.0] | 92.0 | [76.1, 127.0] | 24.1 | [14.0, 28.0] |
| | | | CBR | 4.8 | [3.0, 6.8] | 5.1 | [3.0, 7.1] | 8.4 | [5.7, 10.9] | 1.9 | [1.3, 2.6] | 386.0 | [203.0, 471.0] | 107.8 | [92.2, 139.1] | 24.4 | [14.0, 28.0] |
| | RNT | NO QCH | NO CBR | 4.8 | [3.1, 6.9] | 5.1 | [3.0, 7.2] | 9.2 | [6.4, 12.8] | 2.1 | [1.6, 2.9] | 476.0 | [259.8, 570.8] | 112.7 | [90.0, 138.6] | 5.9 | [4.0, 10.3] |
| | | | CBR | 4.8 | [3.0, 6.8] | 5.0 | [3.0, 7.1] | 9.0 | [7.4, 11.1] | 2.3 | [1.6, 2.8] | 486.0 | [361.0, 559.0] | 122.1 | [94.4, 135.4] | 5.9 | [4.0, 10.3] |
| | | QCH | NO CBR | 4.9 | [3.1, 6.9] | 5.1 | [3.1, 7.2] | 10.0 | [6.1, 12.8] | 2.4 | [1.4, 3.0] | 514.0 | [336.8, 591.0] | 117.7 | [99.2, 135.5] | 5.9 | [4.0, 10.4] |
| | | | CBR | 4.9 | [3.1, 6.9] | 5.1 | [3.1, 7.2] | 9.6 | [6.9, 12.1] | 2.3 | [1.6, 2.7] | 463.5 | [356.0, 555.5] | 126.3 | [99.2, 144.3] | 5.9 | [4.0, 10.2] |

*Tab. S6: Median and IQR of pandemic characteristics for realistic behaviour. This table complements Fig. 3b-d from the main document. In realistic behaviour, we observe that DCT is the main driver of pandemic characteristic reduction, and in combination with RNT, provides strong differentiation for days in quarantine. Using the IQR, we observe that DCT and No RNT yield the best performance group. However, selecting between combinations of QCH and CBR based on the mean and median shows that additional information is required for an adequate selection. The highlighted table rows correspond to the lines from Fig. 3d.*
